# A Biological-Systems-Based Analyses Using Proteomic and Metabolic Network Inference Reveals Mechanistic Insights into Hepatic Lipid Accumulation: An IMI-DIRECT Study

**DOI:** 10.1101/2025.06.02.25328773

**Authors:** Natalie N. Atabaki, Daniel E. Coral, Hugo Pomares-Millan, Kieran Smith, Harry H. Behjat, Robert W. Koivula, Andrea Tura, Hamish Miller, Katherine Pinnick, Leandro Agudelo, Kristine H. Allin, Andrew A. Brown, Elizaveta Chabanova, Piotr J Chmura, Ulrik P. Jacobsen, Adem Y. Dawed, Petra J.M. Elders, Juan J. Fernandez-Tajes, Ian M. Forgie, Mark Haid, Tue H. Hansen, Elizaveta L. Hansen, Angus G. Jones, Tarja Kokkola, Sebastian Kalamajski, Anubha Mahajan, Timothy J. McDonald, Donna McEvoy, Mirthe Muilwijk, Konstantinos D. Tsirigos, Jagadish Vangipurapu, Sabine van Oort, Henrik Vestergaard, Jerzy Adamski, Joline W. Beulens, Søren Brunak, Emmanouil T. Dermitzakis, Giuseppe N. Giordano, Ramneek Gupta, Torben Hansen, Leen t Hart, Andrew T. Hattersley, Leanne Hodson, Markku Laakso, Ruth J.F. Loos, Jordi Merino, Mattias Ohlsson, Oluf Pedersen, Martin Ridderstråle, Hartmut Ruetten, Femke Rutters, Jochen M. Schwenk, Jeremy Tomlinson, Mark Walker, Hanieh Yaghootkar, Fredrik Karpe, Mark I McCarthy, Elizabeth Louise Thomas, Jimmy D. Bell, Andrea Mari, Imre Pavo, Ewan R. Pearson, Ana Viñuela, Paul W. Franks

## Abstract

**Objective:** To delineate organ-specific and systemic drivers of metabolic dysfunction-associated steatotic liver disease (MASLD), we applied integrative causal inference across clinical, imaging, and proteomic domains in individuals with and without type 2 diabetes (T2D).

**Research Design and Methods:** We used Bayesian network analyses to quantify causal pathways linking adipose distribution, glycemia, and insulin dynamics with fatty liver using data from the IMI-DIRECT prospective cohort study. Measurements were made of glucose and insulin dynamics (using frequently-sampled metabolic challenge tests), MRI-derived abdominal and liver fat content, serological biomarkers, and Olink plasma proteomics from 331 adults with new-onset T2D and 964 adults free from diabetes at enrolment. The common protocols used in these two cohorts provided the opportunity for replication analyses to be performed. When the direction of the effect could not be determined with high probability through Bayesian networks, complementary two-sample Mendelian randomization (MR) was employed.

**Results:** High basal insulin secretion rate (BasalISR) was identified as the primary causal driver of liver fat accumulation in both diabetes and non-diabetes. Excess visceral adipose tissue (VAT) was bidirectionally associated with liver fat, indicating a self-reinforcing metabolic loop. Basal insulin clearance (Clinsb) worsened as a consequence of liver fat accumulation to a greater degree before the onset of T2D. Out of 446 analysed proteins, 34 mapped to these metabolic networks and 27 were identified in the non-diabetes network, 18 in the diabetes network, and 11 were common between the two networks. Key proteins directly associated with liver fat included GUSB, ALDH1A1, LPL, IGFBP1/2, CTSD, HMOX1, FGF21, AGRP, and ACE2. Sex-stratified analyses revealed distinct proteomic drivers: GUSB and LEP were most predictive of liver fat in females and males, respectively.

**Conclusions:** Basal insulin hypersecretion is a modifiable, causal driver of MASLD, particularly prior to glycaemic decompensation. Our findings highlight a multifactorial, sex-and disease-stage–specific proteo-metabolic architecture of hepatic steatosis. Proteins such as GUSB, ALDH1A1, LPL, and IGFBPs warrant further investigation as potential biomarkers or therapeutic targets for MASLD prevention and treatment.

## Introduction

About a third of the global adult population is estimated to have metabolic dysfunction-associated steatotic liver disease (MASLD), which is predicted to increase markedly in the coming decades [1]. In people with type 2 diabetes (T2D), the prevalence of MASLD is about 67% and rising [2]. Although the epidemiological association between intrahepatic lipid accumulation and T2D is well described, its underlying metabolic and proteomic features remain poorly understood, hindering the prevention and treatment of MASLD [3].

Hyperinsulinemia, especially excessive early-phase insulin secretion, is a common prelude to T2D and MASLD [4, 5] and the hyper-secretion of insulin often seen in people with T2D is exasperated when MASLD co-occurs [6]. When adipocyte lipid storage capacity is exceeded, ectopic lipid accumulation occurs, with the liver being a major sink for excess triacylglycerol.

An over-abundance of metabolic substrates (glucose and fatty acids) drives liver-specific insulin resistance and corresponding *de novo* hepatic glucose and lipid production. This degenerative cycle leads to systemic elevation in glucose and lipids – major risk factors for T2D [7–10].

Understanding the plasma protein network and its relationship with glucose and insulin dynamics and adipose distribution may elucidate how these metabolic features interact. Recent proteomic studies have explored specific features of MASLD [3, 11–13] Nevertheless, a comprehensive system-wide model is lacking, hindering understanding of the causal drivers and consequences of liver fat accumulation and its effects on glycose and insulin homeostasis. Addressing this gap may inform diagnostic, preventive, and therapeutic strategies for MASLD.

Here, we applied Bayesian network analysis and Mendelian randomization to the comprehensively phenotyped datasets from the IMI-DIRECT cohorts to determine potential causal networks linking plasma proteins, glucose and insulin dynamics with hepatic lipid accumulation.

## MATERIALS AND METHODS

### IMI-DIRECT cohorts and measures

To avoid imputation biases, analyses focused on complete case data from IMI-DIRECT, a multicentre prospective cohort study involving European-ancestry adults diagnosed with T2D (n=331) and those without T2D (n=964). The latter group included participants with normal and prediabetic glycemia (ascertained by glycated haemoglobin A1C (HbA1c), fasting glucose, or 2-hour glucose). All participants provided written informed consent at enrolment, and the study protocol was approved by the regional research ethics committees of each clinical study centre [14, 15].

Frequently-sampled mixed-meal tolerance tests (MMTT) and oral glucose tolerance tests (fsOGTT) were carried out in the diabetes and non-diabetes cohorts, respectively. Insulin secretion estimates were derived from C-peptide deconvolution using the method described by Van Cauter et al. [16], expressed as pmol/min per square meter of estimated body surface area. Visceral and subcutaneous fat were assessed using T1-weighted MRI scans, while liver and pancreas fat content were measured using a T2*-based multi-echo technique from MRI scans [17].

Biospecimens from IMI-DIRECT study centres were manually randomized using a mix-shake-distribute procedure and placed into 96-well plates. Proteins in EDTA plasma samples were quantified at SciLifeLab in Stockholm using the Cardiometabolic, Cardiovascular II, Cardiovascular III, Development, and Metabolism panels provided by Olink Proteomics AB (Uppsala, Sweden), following the guidelines for Proximity Extension Assays (PEA) [18]. Further details on the study design and core characteristics are provided elsewhere [14, 15].

### Bayesian network analysis

We employed Bayesian networks to create directed acyclic graphs (DAGs) that visually depict potential causal connections and interactions between clinical and protein features, using established methods. In these graphs, each variable is represented as a node, and the arcs connecting them suggest possible cause-and-effect relationships.

The network’s skeleton was constructed using the Semi-Interleaved HITON-PC (SI-HITON-PC) algorithm. SI-HITON-PC is a constraint-based algorithm that performs conditional correlation-based independence tests (p-value ≤ 0.01)[19]. This process selects protein features and determines the presence of arcs between clinical and proteomic nodes. The algorithm identifies the Markov blanket for each node, which is the minimal set of nodes that makes the node conditionally independent of other nodes in the network. Directional arcs were then assigned using a hill-climbing algorithm, which orients the arcs based on causal Bayesian theorem while avoiding cycles in the DAG.

These steps were carried out through a 10-fold cross-validation process, aggregating predictive and posterior correlation scores from multiple runs of the model. To create a robust structure that resists network perturbations, we used model averaging, resulting in an averaged network based on the frequency of observed potential arcs across all networks. The strength of each arc was determined by how frequently it appears in the model; the probability of the assigned direction was also calculated. The purpose of these analyses is to determine the probability of true-positive discovery and causal effect directionality. To ensure the networks function appropriately, we inverse-normalized all numeric variables to fit a Gaussian Bayesian network with a multivariate normal distribution.

### Mendelian randomization analysis

Following the construction of the Bayesian networks, we performed bi-directional two-sample Mendelian randomization (MR) analysis [20] on connections between proteins and clinical variables that were strongly associated but with low directional probability (suggesting bidirectional causal associations).

For associations between proteins and clinical variables, we utilized the SMR (Summary-based Mendelian randomization) and HEIDI (Heterogeneity in Dependent Instruments) methodologies [21]. We applied the top pQTL genetic loci as the instrument *cis*-pQTLs within a ±1 MB window of the gene encoding the target protein. HEIDI analysis was conducted to assess the heterogeneity of each lead SNP estimate relative to those in linkage disequilibrium (LD), helping distinguish biological pleiotropic effects (p-value_Heidi > 0.01) from those driven solely by LD.

For reciprocal associations between clinical variables and proteins, we employed the inverse variance weighted (IVW) method as the primary MR analysis along with MR-Egger. We considered MR findings to be statistically significant if the causal association amongst IVW and MR Egger was directionally concordant and the causal association for IVW passed a false discovery rate (FDR) corrected threshold (p-value < 7e^-4^) for multiple testing (i.e., 0.05/(34×2)). To detect and account for directional pleiotropy, the MR-Egger intercept test was conducted (p-value > 0.05). The MR analysis utilized the latest summary statistics from publicly available databases, detailed in Table S1.

Statistical analyses were undertaken using *R* software version 4.2.1. The Bayesian networks were constructed using the *bnlearn* package [22], and the MR analysis was performed using *TwoSampleMR* [23]. Bayesian networks were visualized using *Cytoscape* software version

3.10.3 [24]. The scripts used can be found at https://github.com/Natalie-Atabaki/BN. Variables from IMI-DIRECT were adjusted for the centre of origin of the samples (known as the ‘centre effect’), age, and sex (when appropriate) in a linear model, including each variable per model. For proteins, in addition to biological adjustments, plate adjustments were made. Residuals were extracted from these models and rank normalized to fulfil the Gaussian Bayesian network assumption.

## Results

Within the IMI-DIRECT cohorts, 331 (with T2D) and 964 (without T2D) participants had the required clinical (n=19) and proteomic (n=468) variables for a complete case analysis. An overview of IMI-DIRECT participant characteristics is shown in Table S1 for the non-diabetes and diabetes cohorts. These clinical variables were initially selected based on their association with hepatic lipid accumulation within the IMI-DIRECT cohorts [25] or from existing literature [7–10, 26]; these include glycaemic, beta-cell function, adipose volume, and hepatic lipid content variables.

### Proteomic feature selection in IMI-DIRECT cohorts

Of 446 analysed Olink plasma proteins, 34 were identified as key components in the metabolic network, including CTRC, PON3, KITLG, APOM, CD4, LDLR, MFGE8, LPL, MATN2, FAM3C, FGF21, TGM2, IGFBP1, LEP, AGRP, ANXA4, ACE2, HMOX1, CHL1, MSMB, NRP1, FST, GUSB, GHRL, ANGPTL4, IGFBP2, CDH5, CTSD, SRCRB4D, FCRL5, WFIKKN2, CD300LG, ALDH1A1, and GH1, with 27 present in the non-diabetes network and 18 in the diabetes network, of which 11 were shared across networks. Functional enrichment analysis revealed that these proteins are associated with several cellular components, including protein-lipid macromolecular complexes, LDL particles, and the endoplasmic reticulum lumen. Their molecular functions were enriched in signalling receptor binding, hormone activity, and growth factor interactions. Pathway analysis showed overrepresentation in biological processes related to lipoprotein metabolism, ghrelin processing, and fat-soluble vitamin metabolism, highlighting their potential roles in metabolic regulation across health and disease states (Figure 1) [27–30].

**Figure 1.**
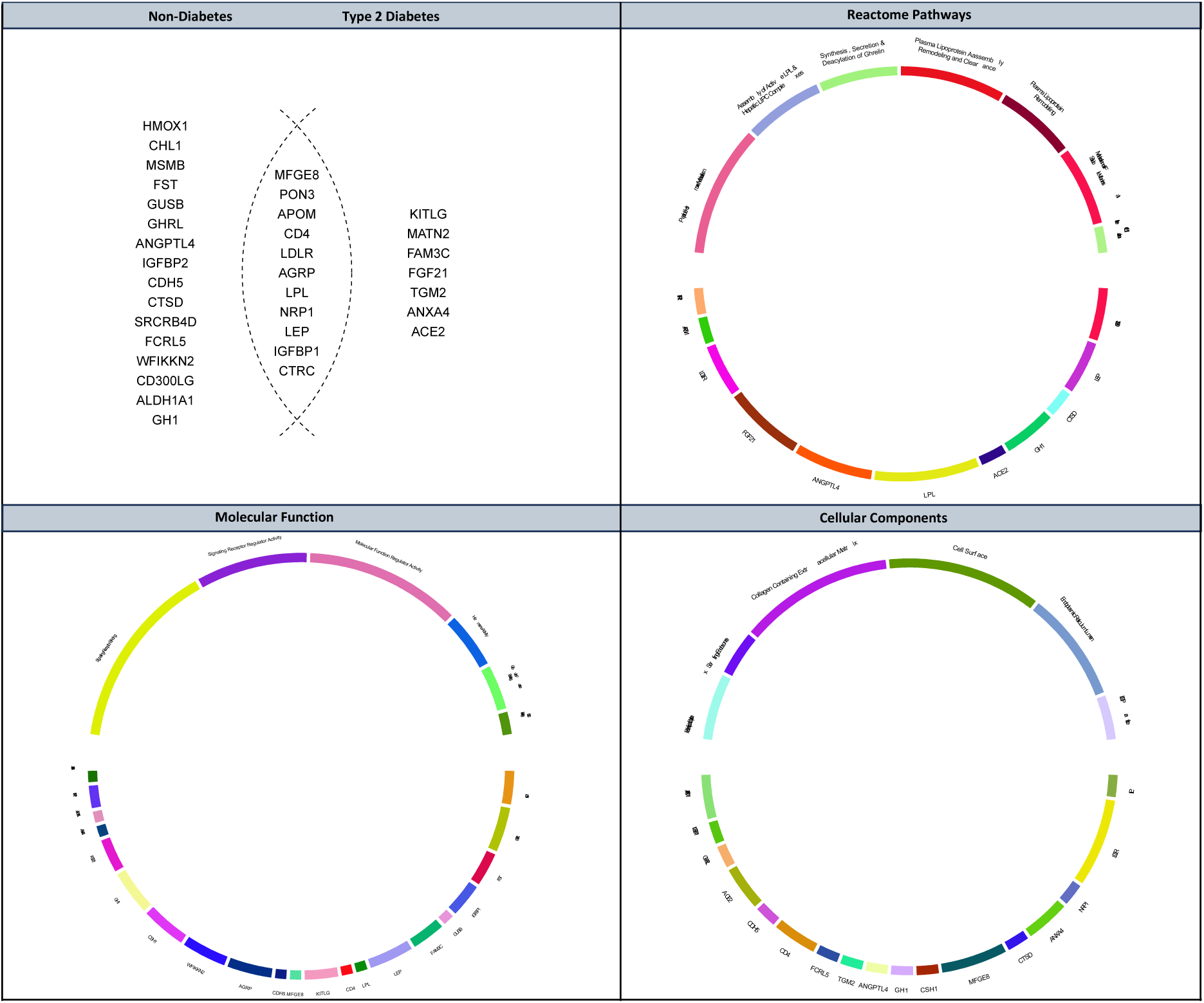
Proteins identified in Bayesian networks and their functional enrichment across metabolic pathways. (Top Left) Venn diagram showing the distribution of 34 proteins identified in the Bayesian networks across non-diabetes and type 2 diabetes cohorts; (Top Right) Reactome pathway enrichment highlighting associated biological processes; (Bottom Left) Molecular function enrichment analysis; (Bottom Right) Cellular component enrichment. The background gene set for the hypergeometric test included 18,731proteining-coding genes.

### Metabolic Bayesian network in IMI-DIRECT Non-Diabetes cohort

To construct the metabolic network, we performed structural and parameter learning using clinical and proteomic features as nodes within the non-diabetic cohort (n=964). To evaluate the network’s fit to the data, we compared the Bayesian information criterion (BIC) scores of the learned network with the BIC density of 1000 randomly generated (null) networks (Figure S1). To ensure robustness and parsimony, we restricted the network to arcs with both strength and directional probabilities ≥0.8. Table 1 outlines the conditional density of each node, along with estimates of the parameters of its upstream (parent) variables, indicating the magnitude of their relationships within the network.

**Table 1.**
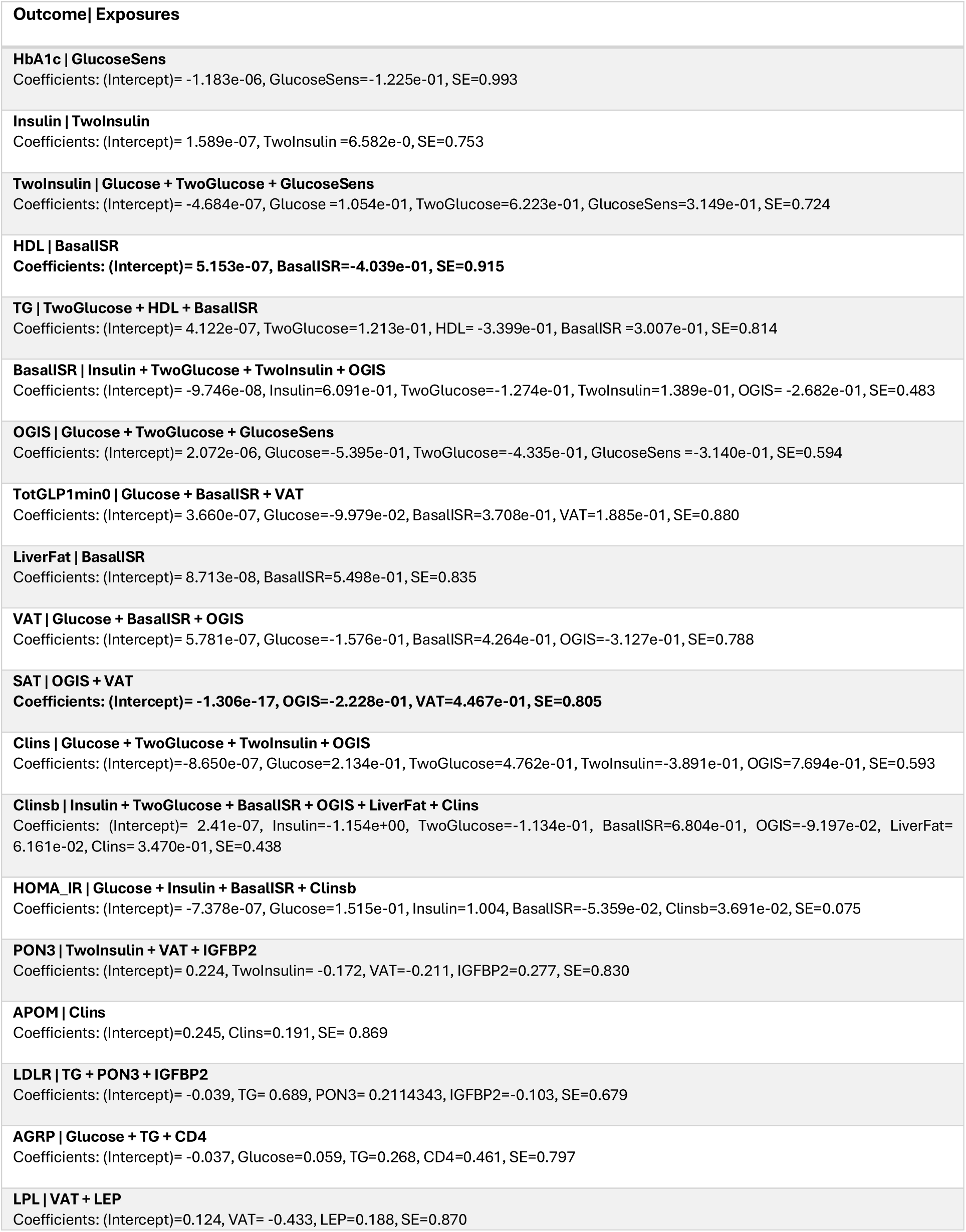
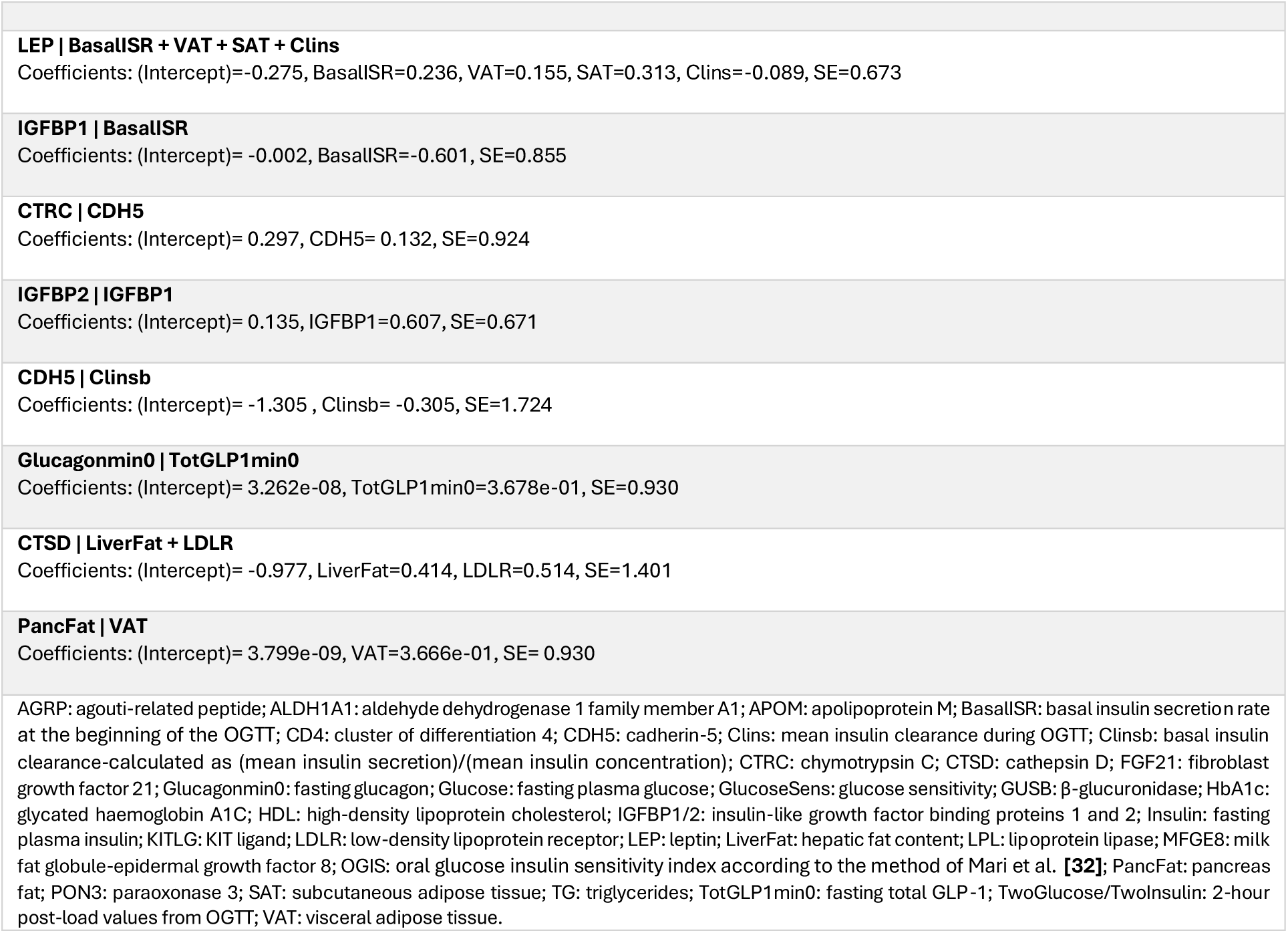
Conditional regression models for each node in the averaged Bayesian network (non-diabetes cohort, n = 964). Each row lists the dependent variable (Outcome) and its set of parent variables (Exposures), along with coefficient estimates and standard error (SE). All variables were inverse-normal transformed.

| **Outcome \| Exposures** |
| --- |
| **HbA1c \| GlucoseSens**<br>Coefficients: (Intercept)= -1.183e-06, GlucoseSens=-1.225e-01, SE=0.993 |
| **Insulin \| TwoInsulin**<br>Coefficients: (Intercept)= 1.589e-07, TwoInsulin =6.582e-0, SE=0.753 |
| **TwoInsulin \| Glucose + TwoGlucose + GlucoseSens**<br>Coefficients: (Intercept)= -4.684e-07, Glucose=1.054e-01, TwoGlucose=6.223e-01, GlucoseSens=3.149e-01, SE=0.724 |
| **HDL \| BasalISR**<br>Coefficients: (Intercept)= 5.153e-07, BasalISR=-4.039e-01, SE=0.915 |
| **TG \| TwoGlucose + HDL + BasalISR**<br>Coefficients: (Intercept)= 4.122e-07, TwoGlucose=1.213e-01, HDL= -3.399e-01, BasalISR =3.007e-01, SE=0.814 |
| **BasalISR \| Insulin + TwoGlucose + TwoInsulin + OGIS**<br>Coefficients: (Intercept)= -9.746e-08, Insulin=6.091e-01, TwoGlucose=-1.274e-01, TwoInsulin=1.389e-01, OGIS= -2.682e-01, SE=0.483 |
| **OGIS \| Glucose + TwoGlucose + GlucoseSens**<br>Coefficients: (Intercept)= 2.072e-06, Glucose=-5.395e-01, TwoGlucose=-4.335e-01, GlucoseSens =-3.140e-01, SE=0.594 |
| **TotGLP1min0 \| Glucose + BasalISR + VAT**<br>Coefficients: (Intercept)= 3.660e-07, Glucose=-9.979e-02, BasalISR=3.708e-01, VAT=1.885e-01, SE=0.880 |
| **LiverFat \| BasalISR**<br>Coefficients: (Intercept)= 8.713e-08, BasalISR=5.498e-01, SE=0.835 |
| **VAT \| Glucose + BasalISR + OGIS**<br>Coefficients: (Intercept)= 5.781e-07, Glucose=-1.576e-01, BasalISR=4.264e-01, OGIS=-3.127e-01, SE=0.788 |
| **SAT \| OGIS + VAT**<br>Coefficients: (Intercept)= -1.306e-17, OGIS=-2.228e-01, VAT=4.467e-01, SE=0.805 |
| **Clins \| Glucose + TwoGlucose + TwoInsulin + OGIS**<br>Coefficients: (Intercept)=-8.650e-07, Glucose=2.134e-01, TwoGlucose=4.762e-01, TwoInsulin=-3.891e-01, OGIS=7.694e-01, SE=0.593 |
| **Clinsb \| Insulin + TwoGlucose + BasalISR + OGIS + LiverFat + Clins**<br>Coefficients: (Intercept)= 2.41e-07, Insulin=-1.154e+00, TwoGlucose=-1.134e-01, BasalISR=6.804e-01, OGIS=-9.197e-02, LiverFat=6.161e-02, Clins=3.470e-01, SE=0.438 |
| **HOMA_IR \| Glucose + Insulin + BasalISR + Clinsb**<br>Coefficients: (Intercept)= -7.378e-07, Glucose=1.515e-01, Insulin=1.004, BasalISR=-5.359e-02, Clinsb=3.691e-02, SE=0.075 |
| **PON3 \| TwoInsulin + VAT + IGFBP2**<br>Coefficients: (Intercept)= 0.224, TwoInsulin= -0.172, VAT=-0.211, IGFBP2=0.277, SE=0.830 |
| **APOM \| Clins**<br>Coefficients: (Intercept)=0.245, Clins=0.191, SE= 0.869 |
| **LDLR \| TG + PON3 + IGFBP2**<br>Coefficients: (Intercept)= -0.039, TG= 0.689, PON3= 0.2114343, IGFBP2=-0.103, SE=0.679 |
| **AGRP \| Glucose + TG + CD4**<br>Coefficients: (Intercept)= -0.037, Glucose=0.059, TG=0.268, CD4=0.461, SE=0.797 |
| **LPL \| VAT + LEP**<br>Coefficients: (Intercept)=0.124, VAT= -0.433, LEP=0.188, SE=0.870 |
| **LEP \| BasalISR + VAT + SAT + Clins**<br>Coefficients: (Intercept)=-0.275, BasalISR=0.236, VAT=0.155, SAT=0.313, Clins=-0.089, SE=0.673 |
| **IGFBP1 \| BasalISR**<br>Coefficients: (Intercept)= -0.002, BasalISR=-0.601, SE=0.855 |
| **CTRC \| CDH5**<br>Coefficients: (Intercept)= 0.297, CDH5= 0.132, SE=0.924 |
| **IGFBP2 \| IGFBP1**<br>Coefficients: (Intercept)= 0.135, IGFBP1=0.607, SE=0.671 |
| **CDH5 \| Clinsb**<br>Coefficients: (Intercept)= -1.305 , Clinsb= -0.305, SE=1.724 |
| **Glucagonmin0 \| TotGLP1min0**<br>Coefficients: (Intercept)= 3.262e-08, TotGLP1min0=3.678e-01, SE=0.930 |
| **CTSD \| LiverFat + LDLR**<br>Coefficients: (Intercept)= -0.977, LiverFat=0.414, LDLR=0.514, SE=1.401 |
| **PancFat \| VAT**<br>Coefficients: (Intercept)= 3.799e-09, VAT=3.666e-01, SE= 0.930 |
| AGRP: agouti-related peptide; ALDH1A1: aldehyde dehydrogenase 1 family member A1; APOM: apolipoprotein M; BasalISR: basal insulin secretion rate at the beginning of the OGTT; CD4: cluster of differentiation 4; CDH5: cadherin-5; Clins: mean insulin clearance during OGTT; Clinsb: basal insulin clearance-calculated as (mean insulin secretion)/(mean insulin concentration); CTRC: chymotrypsin C; CTSD: cathepsin D; FGF21: fibroblast growth factor 21; Glucagonmin0: fasting glucagon; Glucose: fasting plasma glucose; GlucoseSens: glucose sensitivity; GUSB: β-glucuronidase; HbA1c: glycated haemoglobin A1C; HDL: high-density lipoprotein cholesterol; IGFBP1/2: insulin-like growth factor binding proteins 1 and 2; Insulin: fasting plasma insulin; KITLG: KIT ligand; LDLR: low-density lipoprotein receptor; LEP: leptin; LiverFat: hepatic fat content; LPL: lipoprotein lipase; MFGE8: milk fat globule-epidermal growth factor 8; OGIS: oral glucose insulin sensitivity index according to the method of Mari et al. [32]; PancFat: pancreas fat; PON3: paraoxonase 3; SAT: subcutaneous adipose tissue; TG: triglycerides; TotGLP1min0: fasting total GLP-1; TwoGlucose/TwoInsulin: 2-hour post-load values from OGTT; VAT: visceral adipose tissue. |

The constructed network (Figure 2) identified basal insulin secretion rate (BasalISR) as the main direct causal determinant of hepatic lipid content. Insulin clearance, calculated from basal values as 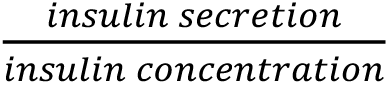 (Clinsb), was identified as a downstream effect of liver fat.

**Figure 2.**
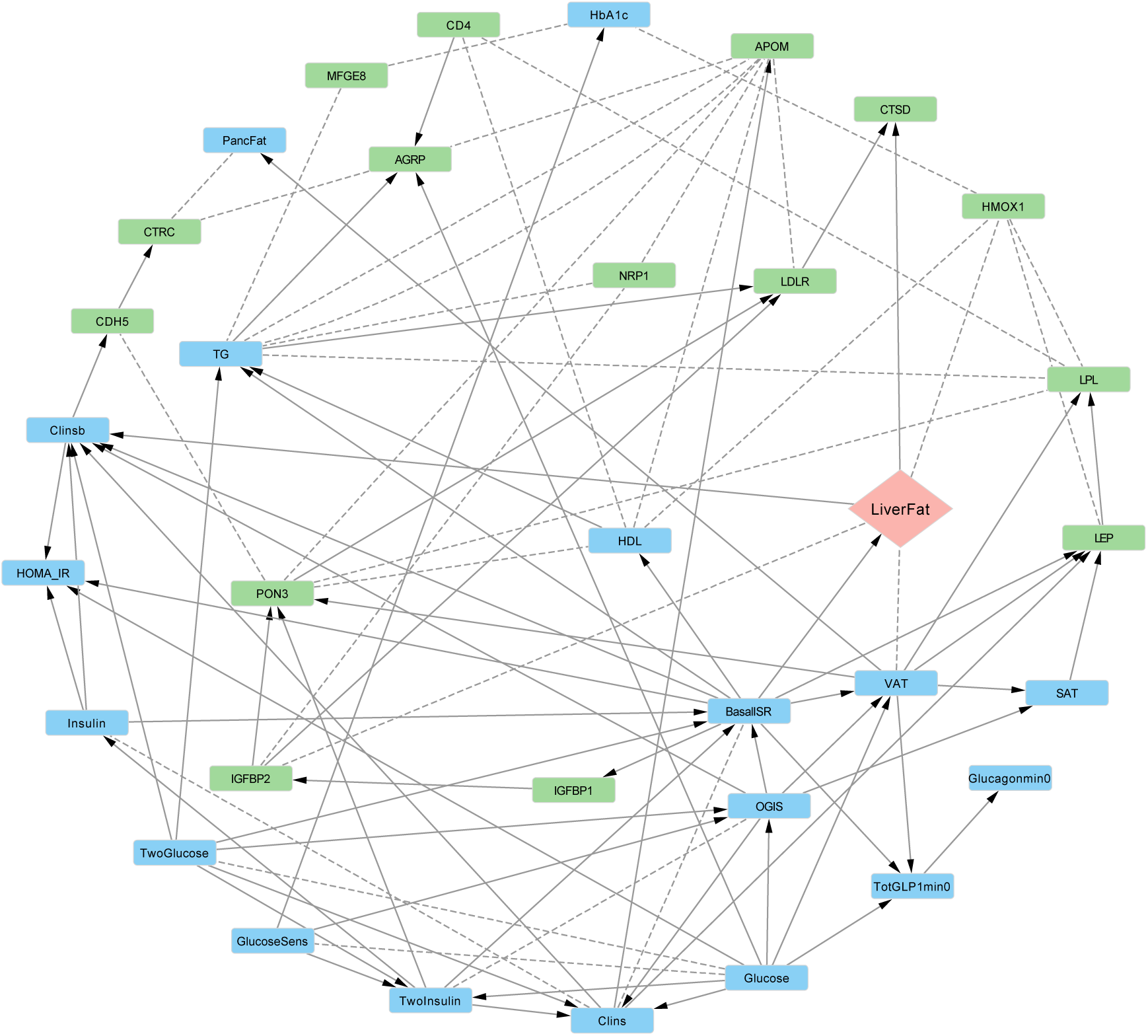
Bayesian network of metabolic and proteomic interactions in the IMI-DIRECT non-diabetes cohort (n = 964). The graph displays directed relationships among clinical and proteomic variables. Nodes are color-coded: blue (clinical/metabolic), green (proteins), peach (liver fat as outcome). Solid arrows represent directed associations with high confidence (strength and direction probability ≥ 0.8), while dashed arrows indicate less confident directionality. AGRP: agouti-related peptide; ALDH1A1: aldehyde dehydrogenase 1 family member A1; APOM: apolipoprotein M; BasalISR: basal insulin secretion rate at the beginning of the OGTT; CD4: cluster of differentiation 4; CDH5: cadherin-5; Clins: mean insulin clearance during OGTT; Clinsb: basal insulin clearance-calculated as (mean insulin secretion)/(mean insulin concentration); CTRC: chymotrypsin C; CTSD: cathepsin D; FGF21: fibroblast growth factor 21; Glucagonmin0: fasting glucagon; Glucose: fasting plasma glucose; GlucoseSens: glucose sensitivity; GUSB: β-glucuronidase; HbA1c: glycated haemoglobin A1C; HDL: high-density lipoprotein cholesterol; IGFBP1/2: insulin-like growth factor binding proteins 1 and 2; Insulin: fasting plasma insulin; KITLG: KIT ligand; LDLR: low-density lipoprotein receptor; LEP: leptin; LiverFat: hepatic fat content; LPL: lipoprotein lipase; MFGE8: milk fat globule-epidermal growth factor 8; OGIS: oral glucose insulin sensitivity index according to the method of Mari et al. [32]; PancFat: pancreas fat; PON3: paraoxonase 3; SAT: subcutaneous adipose tissue; TG: triglycerides; TotGLP1min0: fasting total GLP-1; TwoGlucose/TwoInsulin: 2-hour post-load values from OGTT; VAT: visceral adipose tissue.

BasalISR was the parental node for both liver fat and VAT, leading to fat accumulation in the ectopic and visceral abdominal areas. Given VAT’s strong correlation with liver fat, we hypothesised that BasalISR directly and indirectly (via VAT accumulation) drives liver fat accumulation.

The derived network revealed a direct inverse effect of basal hyperinsulinemia on Insulin-like Growth Factor Binding Protein-1 (IGFBP-1), which then positively affects IGFBP2. Insulin suppresses IGFBP-1 production in the liver, indicating that individuals with higher basal insulin secretion rates have lower IGFBP1 levels owing to this suppressive feedback loop. The network also suggests a direct causal effect of liver fat on CTSD (Cathepsin D) levels, an enzyme involved in protein degradation. CTSD has also been associated with lipid metabolism, inflammation, and hepatic fibrosis in non-alcoholic fatty liver disease (NAFLD) [31]. Higher HMOX1 (Heme Oxygenase 1) levels were strongly associated with increased liver fat, which may be owing to its role in metabolic stress response. This could be either a reaction to liver fat-induced oxidative stress or to the products of heme degradation (e.g., iron) that exacerbate lipid peroxidation, contributing to fat storage and inflammation in the liver. Plasma proteins CTSD, IGFBP2, HMOX1, GUSB (β-glucuronidase), LPL (Lipoprotein Lipase), ALDH1A1 (Aldehyde Dehydrogenase 1 Family Member A1) and AGRP (Agouti-related protein) were directly linked to liver fat at different strength and directional probability levels (Table S1 and Figure S2).

### Metabolic Bayesian network in IMI-DIRECT Diabetes cohort

To identify causal networks specific to individuals with T2D, we conducted Bayesian network analyses within the T2D cohort of the IMI-DIRECT study (n = 331) in a similar manner as before (Figure S3, Table 2, Figure 3). In alignment with the non-diabetes network, BasalISR emerged as a putative upstream driver influencing liver fat accumulation with the FGF21 (fibroblast growth factor 21) protein as a downstream target of liver fat. FGF21, predominantly secreted by the liver, is a key modulator of hepatic glucose and lipid metabolism. Elevated circulating FGF21 levels have been reported in patients with NAFLD [33], possibly reflecting a compensatory response to hepatic lipid accumulation. This response may promote lipolysis, enhance fatty acid oxidation, and improve insulin sensitivity. However, in the context T2D, the effectiveness of this compensatory mechanism may be diminished due to pre-existing metabolic dysregulation.

**Figure 3.**
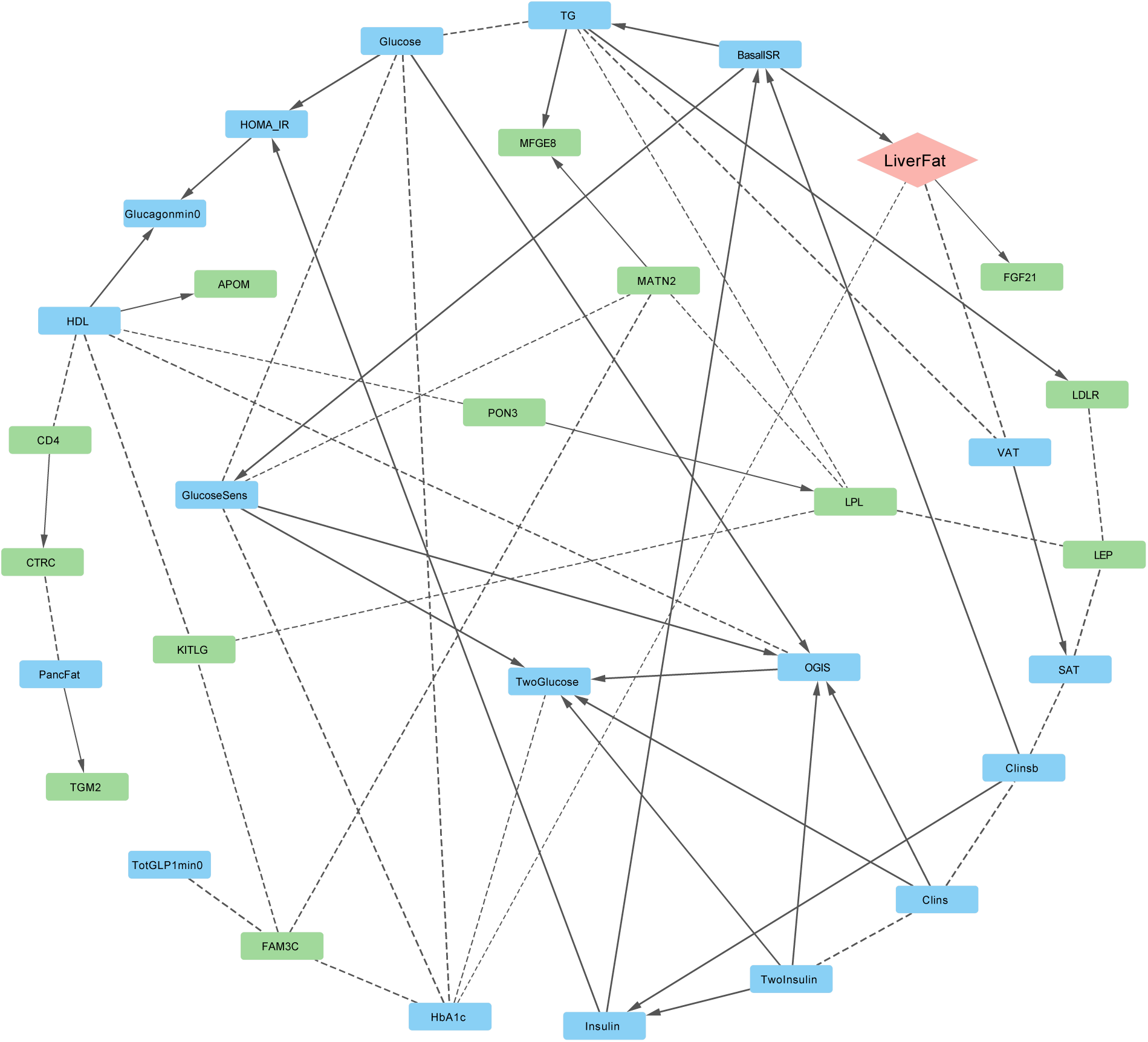
Bayesian network of metabolic and proteomic interactions in the IMI-DIRECT type 2 diabetes cohort (n = 331). The graph displays directed relationships among clinical and proteomic variables. Nodes are color-coded: blue (clinical/metabolic), green (proteins), peach (liver fat as outcome). Solid arrows represent directed associations with high confidence (strength and direction probability ≥ 0.8), while dashed arrows indicate less confident directionality. APOM: apolipoprotein M; BasalISR: basal insulin secretion rate; CD4: cluster of differentiation 4; Clins: mean insulin clearance during OGTT; Clinsb: basal insulin clearance; CTRC: chymotrypsin C; FGF21: fibroblast growth factor 21; Glucagonmin0: fasting glucagon; Glucose: fasting plasma glucose; GlucoseSens: glucose sensitivity; HDL: high-density lipoprotein cholesterol; HOMA_IR: homeostatic model assessment of insulin resistance; Insulin: fasting plasma insulin; LDLR: low-density lipoprotein receptor; LiverFat: hepatic fat content; LPL: lipoprotein lipase; MATN2: matrilin-2; MFGE8: milk fat globule-EGF factor 8; OGIS: oral glucose insulin sensitivity index according to the method of Mari et al. [32]; PancFat: pancreas fat; PON3: paraoxonase 3; SAT: subcutaneous adipose tissue; TG: triglycerides; TGM2: transglutaminase 2; TwoGlucose: 2-hour post-load glucose (OGTT); TwoInsulin: 2-hour post-load insulin (OGTT); VAT: visceral adipose tissue.

**Table 2.**
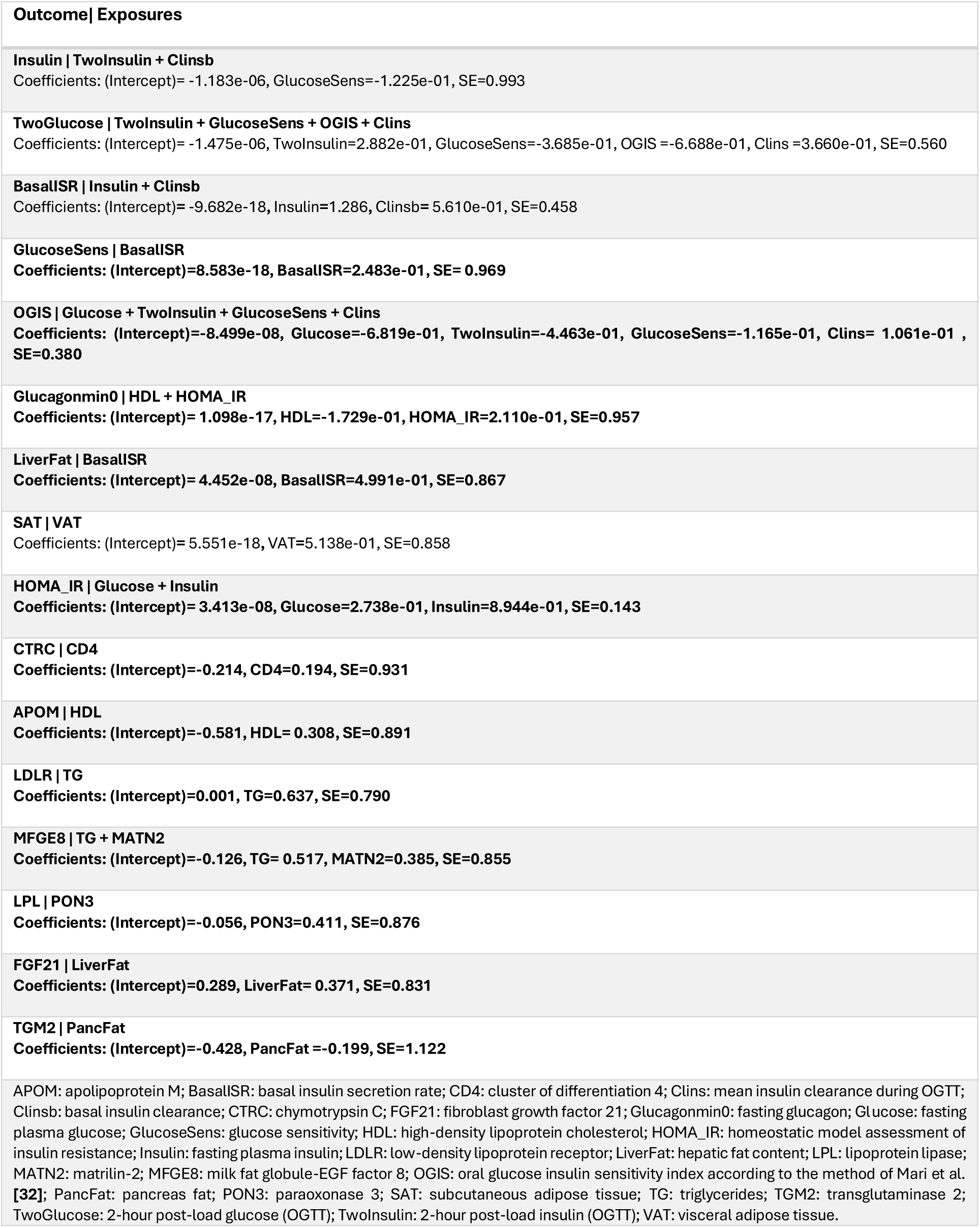
Conditional regression models for each node in the averaged Bayesian network (type 2 diabetes cohort, n = 331). Each row lists the dependent variable (Outcome) and its set of parent variables (Exposures), along with coefficient estimates and standard error (SE). All variables were inverse-normal transformed.

| Outcome \| Exposures |
| --- |
| **Insulin \| TwoInsulin + Clinsb**<br>Coefficients: (Intercept)= -1.183e-06, GlucoseSens=-1.225e-01, SE=0.993 |
| **TwoGlucose \| TwoInsulin + GlucoseSens + OGIS + Clins**<br>Coefficients: (Intercept)= -1.475e-06, TwoInsulin=2.882e-01, GlucoseSens=-3.685e-01, OGIS =-6.688e-01, Clins =3.660e-01, SE=0.560 |
| **BasalISR \| Insulin + Clinsb**<br>Coefficients: (Intercept)= -9.682e-18, Insulin=1.286, Clinsb= 5.610e-01, SE=0.458 |
| **GlucoseSens \| BasalISR**<br>Coefficients: (Intercept)=8.583e-18, BasalISR=2.483e-01, SE= 0.969 |
| **OGIS \| Glucose + TwoInsulin + GlucoseSens + Clins**<br>Coefficients: (Intercept)=-8.499e-08, Glucose=-6.819e-01, TwoInsulin=-4.463e-01, GlucoseSens=-1.165e-01, Clins= 1.061e-01 , SE=0.380 |
| **Glucagonmin0 \| HDL + HOMA_IR**<br>Coefficients: (Intercept)= 1.098e-17, HDL=-1.729e-01, HOMA_IR=2.110e-01, SE=0.957 |
| **LiverFat \| BasalISR**<br>Coefficients: (Intercept)= 4.452e-08, BasalISR=4.991e-01, SE=0.867 |
| **SAT \| VAT**<br>Coefficients: (Intercept)= 5.551e-18, VAT=5.138e-01, SE=0.858 |
| **HOMA_IR \| Glucose + Insulin**<br>Coefficients: (Intercept)= 3.413e-08, Glucose=2.738e-01, Insulin=8.944e-01, SE=0.143 |
| **CTRC \| CD4**<br>Coefficients: (Intercept)=-0.214, CD4=0.194, SE=0.931 |
| **APOM \| HDL**<br>Coefficients: (Intercept)=-0.581, HDL= 0.308, SE=0.891 |
| **LDLR \| TG**<br>Coefficients: (Intercept)=0.001, TG=0.637, SE=0.790 |
| **MFGE8 \| TG + MATN2**<br>Coefficients: (Intercept)=-0.126, TG= 0.517, MATN2=0.385, SE=0.855 |
| **LPL \| PON3**<br>Coefficients: (Intercept)=-0.056, PON3=0.411, SE=0.876 |
| **FGF21 \| LiverFat**<br>Coefficients: (Intercept)=0.289, LiverFat= 0.371, SE=0.831 |
| **TGM2 \| PancFat**<br>Coefficients: (Intercept)=-0.428, PancFat =-0.199, SE=1.122 |
| APOM: apolipoprotein M; BasalISR: basal insulin secretion rate; CD4: cluster of differentiation 4; Clins: mean insulin clearance during OGTT; Clinsb: basal insulin clearance; CTRC: chymotrypsin C; FGF21: fibroblast growth factor 21; Glucagonmin0: fasting glucagon; Glucose: fasting plasma glucose; GlucoseSens: glucose sensitivity; HDL: high-density lipoprotein cholesterol; HOMA_IR: homeostatic model assessment of insulin resistance; Insulin: fasting plasma insulin; LDLR: low-density lipoprotein receptor; LiverFat: hepatic fat content; LPL: lipoprotein lipase; MATN2: matrilin-2; MFGE8: milk fat globule-EGF factor 8; OGIS: oral glucose insulin sensitivity index according to the method of Mari et al. [32]; PancFat: pancreas fat; PON3: paraoxonase 3; SAT: subcutaneous adipose tissue; TG: triglycerides; TGM2: transglutaminase 2; TwoGlucose: 2-hour post-load glucose (OGTT); TwoInsulin: 2-hour post-load insulin (OGTT); VAT: visceral adipose tissue. |

ACE2 (angiotensin-converting enzyme 2) was also directly linked to liver fat, albeit with a weaker association that excluded it from the parsimonious network (Figure S4). As a component of the renin-angiotensin system (RAS), ACE2 mitigates the pro-inflammatory and pro-fibrotic effects of angiotensin II, and its elevation in fatty liver disease may reflect a protective, compensatory adaptation to lipid overload and oxidative stress [34]. Notably, increased hepatic ACE2 expression has been implicated in facilitating SARS-CoV-2 entry, potentially contributing to liver injury and adverse outcomes in individuals with diabetes [35]. Additionally, both VAT and HbA1c were strongly associated with liver fat, although no definitive causal direction could be established (shown as dashed lines in Figure 3). Detailed information of all arcs is reported in Table S1 and Figure S4.

### Complementary directional inference with Mendelian randomization

For some of the arcs, although the strength of the connection was high, the direction of the connection could not be determined with high probability (dashed lines in Figures 2 and 3). This suggests either a bidirectional association or insufficient data to confidently determine causal direction. To investigate this, we performed two-sample MR analysis for connections from clinical variables to the proteins and SMR-HEIDI to study the association from proteins to the clinical traits. The results are reported in Table 3.

**Table 3.**
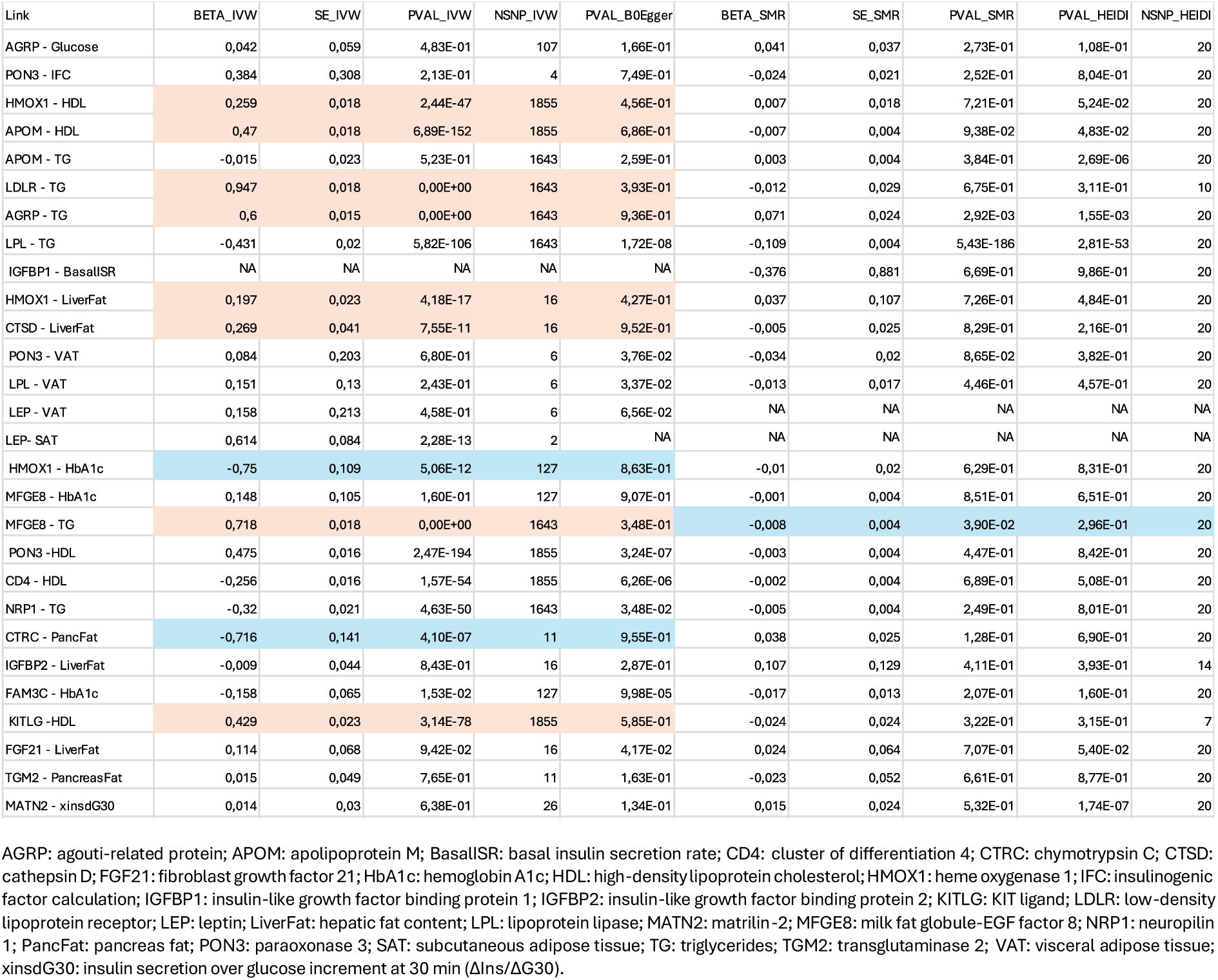
Mendelian Randomization (MR) Analysis to Determine Directionality of Associations Between Clinical Variables and Proteins. This table presents the results of bidirectional MR and SMR-HEIDI analyses used to infer the direction of causal relationships between clinical traits and proteins for which Bayesian networks indicated strong associations but uncertain directionality (Figures 2 and 3). Highlighted rows represent statistically significant findings after correction, with effect estimates, standard errors, and P-values shown for both forward (clinical → protein) and reverse (protein → clinical) directions. Colour shading indicates the direction with stronger evidence: peach for positive beta estimates, and blue for negative beta estimates.

We identified potential positive causal associations from high-density lipoprotein (HDL) to HMOX1, APOM (apolipoprotein M), and KITLG (KIT ligand) proteins. HDL is well-known for its anti-inflammatory and antioxidant properties. The observed association with HMOX1, a stress-response enzyme involved in heme degradation and cytoprotection, may reflect HDL’s role in mitigating oxidative stress through upregulation of HMOX1. APOM, predominantly associated with HDL particles, plays a role in lipid metabolism and endothelial function, so a causal link from HDL aligns with its known role as a carrier protein that influences APOM stability and function [36]. The association with KITLG, a cytokine involved in haematopoiesis and cell survival, may reflect HDL’s regulatory effects on inflammatory and hematopoietic signalling pathways [37]. Triglycerides (TG) also appeared to have positive causal effects on LDLR (low-density lipoprotein receptor) and AGRP (agouti-related peptide) protein levels. Elevated TG levels can lead to changes in lipoprotein metabolism, which may upregulate LDLR expression to enhance lipid clearance from circulation [38]. The link to AGRP, a neuropeptide that promotes appetite and energy intake, may reflect a compensatory mechanism where increased TG levels signal energy abundance, thereby stimulating AGRP expression as part of a homeostatic feedback loop in energy balance.

Liver fat showed a positive causal effect on HMOX1 and CTSD. Increased hepatic lipid accumulation can induce oxidative stress and inflammation, which may trigger upregulation of HMOX1 as a protective response [39]. CTSD, a lysosomal protease involved in protein degradation and lipid metabolism, may be upregulated in response to liver fat to support intracellular lipid turnover and mitigate lipotoxicity [31]. HbA1c and pancreas fat showed negative causal effects on HMOX1 and CTRC (chymotrypsin C), respectively. Chronic hyperglycaemia, characterised here by elevated HbA1c, may suppress HMOX1 through mechanisms involving oxidative damage and altered transcriptional regulation. The negative association between pancreatic fat and CTRC, a pancreatic enzyme that regulates the activation of digestive enzymes, may be explained by fat infiltration inducing lipotoxicity. This, in turn, can stress and impair the function of acinar cells (the primary source of CTRC) potentially reducing its expression or secretion.

MFGE8 (Milk fat globule-EGF factor 8 protein) and TG have bidirectional effects, where TG positively influences MFGE8 and MFGE8 negatively influences TG, suggest a feedback mechanism. Specifically, increased TG levels may upregulate MFGE8 expression to modulate inflammation and phagocytosis of apoptotic cells, which are elevated in metabolic dysfunction [40]. In turn, elevated MFGE8 may enhance lipid clearance or promote anti-inflammatory effects that reduce circulating TG levels, thus helping to restore metabolic balance. MFGE8 has also been implicated in promoting tissue repair and vascular homeostasis, which may be activated in response to lipid-induced damage.

The complementary MR analyses helped assign directionality to ten associations identified through Bayesian network analysis that demonstrated strong associations but where directionality could not be confidently ascertained.

### Bayesian networks and posterior probabilistic inference in IMI-DIRECT

Bayesian networks were constructed to model conditional dependencies and infer potential causal relationships among clinical and proteomic variables in relation to liver fat accumulation. Focusing on liver fat as the outcome of interest, we estimated posterior probabilities by conditioning not only on its direct parent nodes but also individually on each variable in the network. This comprehensive conditioning strategy enabled the identification of both direct and indirect influences on the likelihood of MASLD (defined as liver fat >5%). To facilitate this analysis, all continuous variables were discretized into low, moderate, and high levels using Hartemink’s method [41] (Table S1).

In the non-diabetes network, the baseline probability of MASLD was 34%, which rose to 71% after conditioning on high BasalISR (Figure 4). Proteomic data further revealed distinct modulators of MASLD risk (Figure 5). In this group, the greatest increases (∼30%) in MASLD probability were observed after conditioning on high levels of GUSB and ALDH1A1. GUSB is involved in lysosomal degradation and has been implicated in inflammation and obesity-related metabolic dysfunction [3, 42]; its upregulation may signal hepatic stress or immune activation. NAFLD-associated genetic variants have also been reported to correlate with plasma GUSB levels [3]. ALDH1A1 is a key enzyme in retinoid metabolism and lipid homeostasis and has been linked to hepatic lipid accumulation through its regulation of PPAR signalling [43, 44]. Additionally, low levels of IGFBP1 and LPL were associated with increased MASLD risk (∼15%). IGFBP1 is negatively regulated by insulin and is typically suppressed in insulin-resistant states, serving as a surrogate marker of hepatic insulin action [45]. LPL hydrolyzes circulating triglycerides; reduced activity can impair lipid clearance, promoting ectopic fat deposition in the live [46].

**Figure 4.**
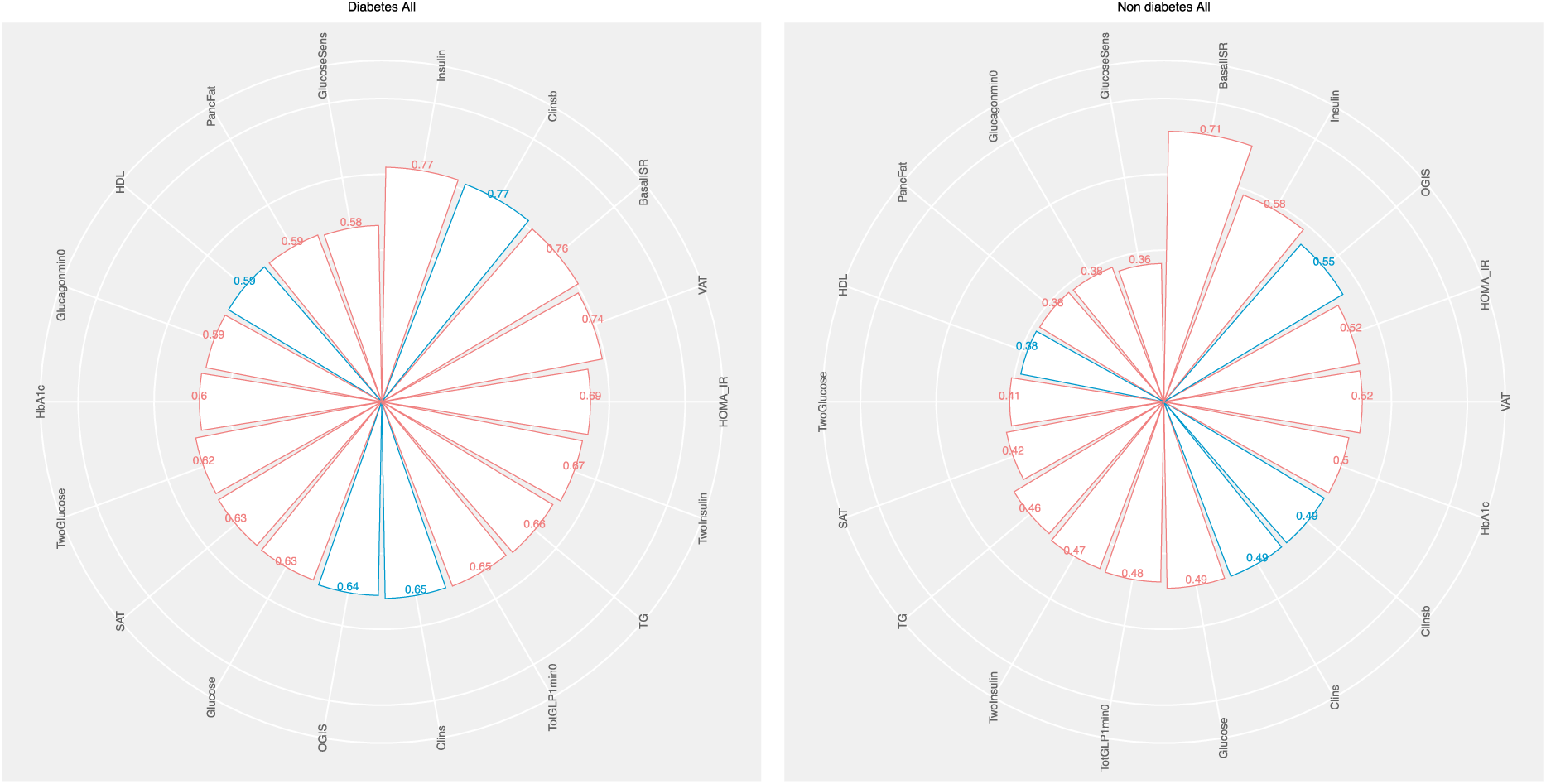
Posterior probabilities of MASLD based on clinical predictors across IMI-DIRECT diabetes and non-diabetes cohorts. Radar plots display the posterior probability of MASLD (metabolic dysfunction-associated steatotic liver disease, defined as liver fat >5%) after conditioning on individual clinical variables in (Left) individuals with T2D (type 2 diabetes) and (Right) those without diabetes. Bars represent the conditional probability of MASLD given high (red bars) or low (blue bars) levels of each variable. MASLD: metabolic dysfunction-associated steatotic liver disease; T2D: type 2 diabetes; BasalISR: basal insulin secretion rate; VAT: visceral adipose tissue; SAT: subcutaneous adipose tissue; OGIS: oral glucose insulin sensitivity index; HOMA_IR: homeostatic model assessment of insulin resistance; Clinsb: basal insulin clearance; Clins: dynamic insulin clearance; TG: triglycerides; HDL: high-density lipoprotein cholesterol; HbA1c: glycated haemoglobin A1c; GLP1: glucagon-like peptide 1; OGTT: oral glucose tolerance test.

**Figure 5.**
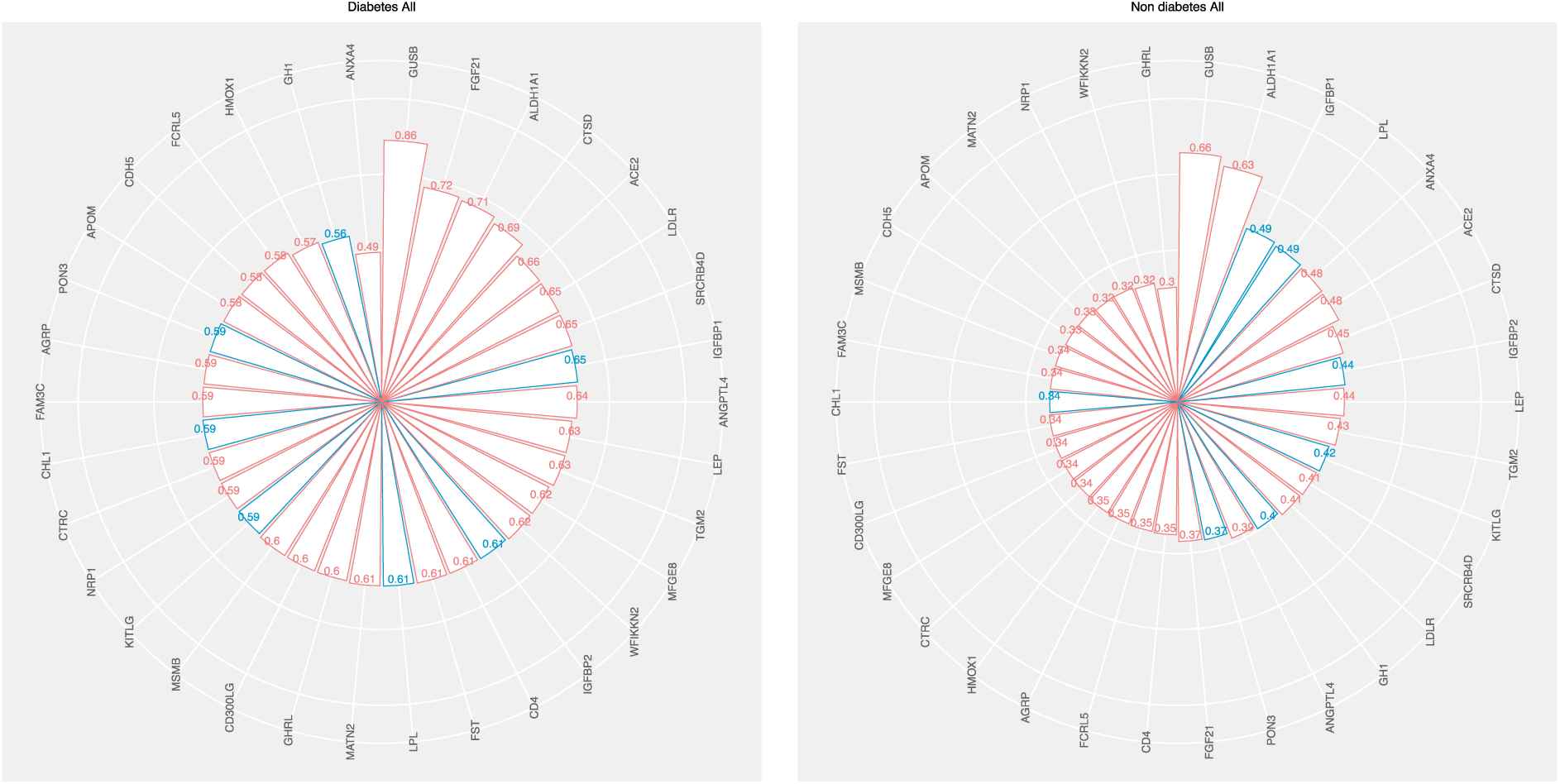
Posterior probabilities of MASLD based on proteomic predictors in IMI-DIRECT diabetes and non-diabetes cohorts. Radar plots display the conditional probability of MASLD after conditioning on individual proteomic variables in (Left) T2D and (Right) non-diabetes cohorts. Bars indicate high (red) or low (blue) levels of the corresponding protein. MASLD: metabolic dysfunction-associated steatotic liver disease; T2D: type 2 diabetes; GUSB: β-glucuronidase; LEP: leptin; ALDH1A1: aldehyde dehydrogenase 1 family member A1; CTSD: cathepsin D; FGF21: fibroblast growth factor 21; IGFBP1/2: insulin-like growth factor binding proteins 1 and 2; LDLR: low-density lipoprotein receptor; MFGE8: milk fat globule-EGF factor 8; AGRP: agouti-related peptide; KITLG: KIT ligand; ACE2: angiotensin-converting enzyme 2; APOM: apolipoprotein M; CD4: cluster of differentiation 4; PON3: paraoxonase 3; TGM2: transglutaminase 2; MATN2: matrilin-2.

In the diabetes network, the unconditioned MASLD probability was 58%, with similar increases (∼20%) observed after conditioning on high fasting insulin, BasalISR, or VAT, and low Clinsb, reinforcing the roles of hyperinsulinemia, insulin resistance, and visceral adiposity in hepatic steatosis. In this network, GUSB again demonstrated the strongest association (∼30%), further supporting its potential role in disease progression. High levels of FGF21, ALDH1A1, and CTSD (∼15%) also elevated MASLD probability. Across both the non-diabetes and diabetes networks, the most informative combination of markers predicting the highest MASLD risk included elevated BasalISR, VAT, GUSB, and ALDH1A1, alongside low levels of LPL and IGFBP1, highlighting a coordinated disruption of pathways related to insulin resistance, inflammation, and lipid metabolism.

Sex-stratified analyses revealed subgroup-specific patterns (Figures S5 and S6). Among non-diabetic males (n = 793), ALDH1A1 and LEP (leptin) showed the strongest effects, while GUSB was most influential in non-diabetic females (n = 171). LEP is central to energy homeostasis and hepatic fat oxidation; its dysregulation may impair hepatic lipid handling, particularly in males [47]. Elevated LEP levels in fatty liver may indicate hepatic LEP resistance, where LEP fails to stimulate lipid turnover. The greater influence of GUSB in females is consistent with prior findings suggesting sex-specific regulation, potentially mediated by oestrogen and body composition. GUSB activity has been reported to be higher in overweight females (but not males), possibly reflecting hormonal fluctuations and systemic low-grade inflammation during reproductive transitions [48]. In the T2D cohort (n = 193 males, n = 138 females), LEP remained the most influential proteomic marker in males, while GUSB remained dominant in females. Among clinical variables, BasalISR and VAT were the strongest predictors of MASLD in females, underscoring the pivotal role of insulin secretion and visceral adiposity in hepatic lipid accumulation. In males, BasalISR was most predictive in the non-diabetic state, whereas subcutaneous adipose tissue (SAT) had greater impact in diabetes, highlighting sex-and disease-stage–specific differences in adipose distribution and metabolic regulation.

## Discussion

This analysis focused on determining causal mechanisms underlying liver fat accumulation in adults with and without T2D using comprehensive clinical, imaging, and proteomic phenotyping in the IMI-DIRECT cohorts. We describe complex interactions between insulin dynamics, adipose distribution, and specific circulating proteins in MASLD risk. Several key findings replicated across cohorts, with some effects being specific to sex and disease state.

One of the key replicated findings is that BasalISR is a primary causal driver of liver fat accumulation, with a stronger effect in individuals without diabetes, providing evidence that basal hyperinsulinemia is a causal driver of hepatic fat accumulation, rather than simply co-occurring or as a downstream effect. This aligns with prior mechanistic evidence indicating that insulin influences hepatic lipid metabolism via *de novo* lipogenesis, more through hormonal regulation (insulin levels) than because of substrate availability (glucose levels) [49]. The close association between excessive insulin secretion and metabolic dyshomeostasis has been shown in several studies, although the causal dynamics between hyperinsulinemia and metabolic dysfunction have hitherto lacked strong evidence [5]. There is, though, an established causal association of excess adiposity on basal insulin secretion rate, even in the absence of insulin resistance, with diet-restricted weight loss lowering insulin secretion rate without changing insulin sensitivity [50].

Our results concur with prior research showing that hyperinsulinemia is a modifiable risk factor for metabolic deterioration, even in people with normoglycemia, and that small changes in insulin levels can affect lipid accumulation and adipogenesis independent of glucose homeostasis [51, 52]. The directionality of relationships between insulin secretion, insulin resistance, and liver fat is complex [53], with evidence supporting both insulin resistance driving hyperinsulinemia [54] and vice versa [49, 55]. In our Bayesian networks, an arc from BasalISR to HOMA-IR and from OGIS (insulin sensitivity) to BasalISR was observed, reflecting the complexity of these effects.

The Bayesian networks also uncovered a bidirectional relationship between liver fat and visceral fat, suggesting a self-perpetuating cycle of fat redistribution and metabolic stress. In the T2D cohort, while BasalISR remained important, VAT exerted similar causal influence on liver fat, likely reflecting the shift in disease drivers as insulin secretion capacity diminishes. Cross-talk between VAT and the liver, mediated by free fatty acids entering the portal circulation, likely underpins this relationship [7]. Additionally, a negative association between fasting insulin clearance and liver fat was observed, suggesting hepatic steatosis may impair insulin catabolism, exacerbating systemic hyperinsulinemia. Sex-specific differences in adipose distribution emerged: visceral fat was the strongest predictor of MASLD in females, while subcutaneous fat had a stronger influence in males with diabetes. These findings highlight the need for sex-and disease-stage-specific interventions that consider differences in fat compartment function and metabolic regulation.

Proteomic analyses revealed 34 proteins associated with liver fat in the Bayesian networks, with 11 replicating across both diabetes and non-diabetes cohorts. Proteins such as GUSB, ALDH1A1, LPL, IGFBP1/2, CTSD, HMOX1, FGF21, AGRP, and LEP played central roles in modulating MASLD risk. GUSB, a lysosomal enzyme linked to inflammation and cellular stress, showed the strongest association with MASLD in both cohorts and was particularly important in females. ALDH1A1, a key regulator of retinoid metabolism and lipid homeostasis, had strong effects in males, and was consistently associated with MASLD risk across metabolic states. LPL and IGFBP1, inversely associated with MASLD, reflect disturbances in lipid clearance and hepatic insulin sensitivity, respectively. Their depletion likely contributes to ectopic fat accumulation and systemic insulin resistance. CTSD and HMOX1, both enzymes involved in oxidative and lysosomal stress responses, were upregulated in relation to liver fat, suggesting a compensatory reaction to lipotoxicity. FGF21, particularly in T2D, appeared as a stress-induced hepatokine, reinforcing its role as a compensatory factor in advanced metabolic dysfunction.

Sex-stratified Bayesian inference further revealed that LEP and ALDH1A1 were more influential in non-diabetic men, while GUSB dominated in females, underscoring sex-specific immune-metabolic mechanisms of liver fat regulation. By estimating posterior probabilities conditioned on single variables and combinations of key markers, we were able to quantify how specific features modify MASLD risk. The greatest increases in MASLD probability were observed following conditioning on combinations of high BasalISR, VAT, GUSB, ALDH1A1and low IGFBP1 and LPL, demonstrating the additive influence of insulin dynamics and proteomic factors. These probabilistic models highlight the clinical utility of combining hormonal and proteomic data to stratify populations by MASLD risk.

Where causal effect directionality was unclear in the Bayesian networks, we applied bidirectional Mendelian randomization. This analysis validated ten associations and helped assign directionality: HDL to HMOX1, APOM, KITLG: reflecting HDL’s antioxidant and signalling roles. TG to LDLR, AGRP: suggesting lipid-induced shifts in receptor expression and appetite regulation. Liver fat to HMOX1, CTSD: supporting a model where hepatic lipid accumulation triggers oxidative and lysosomal stress responses. HbA1c and pancreas fat lowering HMOX1 and CTRC: indicating chronic hyperglycaemia and fat infiltration suppress protective or digestive pathways. A feedback loop between TG and MFGE8 was observed, suggesting reciprocal regulation between lipids and anti-inflammatory responses. These findings enhance the interpretability of our network results and help delineate causal vs. consequential biological processes in MASLD.

The integration of multiple phenotypic layers (including clinical, imaging, metabolic, and proteomic data) within the causal modelling framework deployed here provides robust inference of directionality and mechanistic insight into complex metabolic interactions. Despite these strengths, we should also highlight several key limitations: The cohort consisted solely of individuals of European ancestry, limiting generalizability. Furthermore, MR assumptions may not hold for all proteins due to pleiotropy or weak instruments, and Bayesian inference depends on the completeness and quality of included variables and assumptions. Although several key findings were replicated, it is important to stress that the differences between the two cohorts (i.e., one is of people without and the other with diabetes at enrolment) means that some or all the findings that did not replicate may simply reflect disease-state-specific effects. Disentangling this is challenging, as no comparable cohorts are available for replication.

In summary, this study provides a comprehensive systems-level analysis of MASLD pathogenesis. We show that basal insulin hypersecretion is a modifiable, causal driver of liver fat accumulation, particularly in individuals without diabetes. Through probabilistic inference and genetic validation, we demonstrate that insulin dynamics, visceral fat, and circulating proteins such as GUSB and ALDH1A1 interact in sex-and disease-specific ways to influence liver fat risk. These insights can inform precision medicine approaches to MASLD prevention, including targeted metabolic modulation in high-risk individuals and the potential use of plasma proteomics for risk stratification. Future research should examine the longitudinal impact of modifying these pathways and extend analyses to more diverse populations.

## ETHICS STATEMENTS

Approval for the study protocol was obtained from each of the regional research ethics review boards separately (Lund, Sweden: 20130312105459927, Copenhagen, Denmark: H-1-2012-166 and H-1-2012-100, Amsterdam, Netherlands: NL40099.029.12, Newcastle, Dundee and Exeter, UK: 12/NE/0132). All participants provided written informed consent at enrolment. The research conformed to the ethical principles for medical research involving human participants outlined in the Declaration of Helsinki.

## Data Availability

All data produced in the present study are available upon reasonable request to the authors

## ACKNOWLEDGEMENTS

The work leading to this publication has received support from the Innovative Medicines Initiative Joint. Undertaking under grant agreement n°115317 (DIRECT, https://directdiabetes.org/), resources of which are composed of financial contribution from the European Union’s Seventh Framework Programme (FP7/2007-2013) and EFPIA companies’ in-kind contribution. We thank all the participants and study centre staff in IMI DIRECT for their contribution to the study. NNA was funded by the Swedish Research Council (VR - Avtals-ID: 2021-06714_3) for her postdoctoral fellowship (2022-2025) and acknowledges support from the Henning och Johan Throne-Holsts Foundation. NNA, RJFL and JM are supported by the Novo Nordisk Foundation grant NNF23SA0084103. R.J.F.L. is supported by grants from the National Institutes of Health (R01DK107786; R01DK110113; R01HG010297; R01HL142302; R01DK075787; R01HL156991; U01HG011723; R01DK123019; R01HL151152; R01HL158884), the Danish National Research Fund (DNRF161) and the Novo Nordisk Foundation (NNF20OC0059313). JM acknowledges awards from EFSD/Novo Nordisk Foundation Future Leaders Award (no. 0094134) and the European Union (HORIZON-EIC-2023-PATHFINDERCHALLENGES-01-101161509). Views and opinions expressed are, however, those of the author(s) only and do not necessarily reflect those of the European Union or European Innovation Council and SMEs Executive Agency (EISMEA). Neither the European Union nor the granting authority can be held responsible for them. PWF was supported by grants from the Swedish Research Council (#2019-01348), Swedish Foundation for Strategic Research (LUDC-IRC, 15-0067), and the European Commission (ERC-CoG_NASCENT - 681742).

---

**Figure S1.**
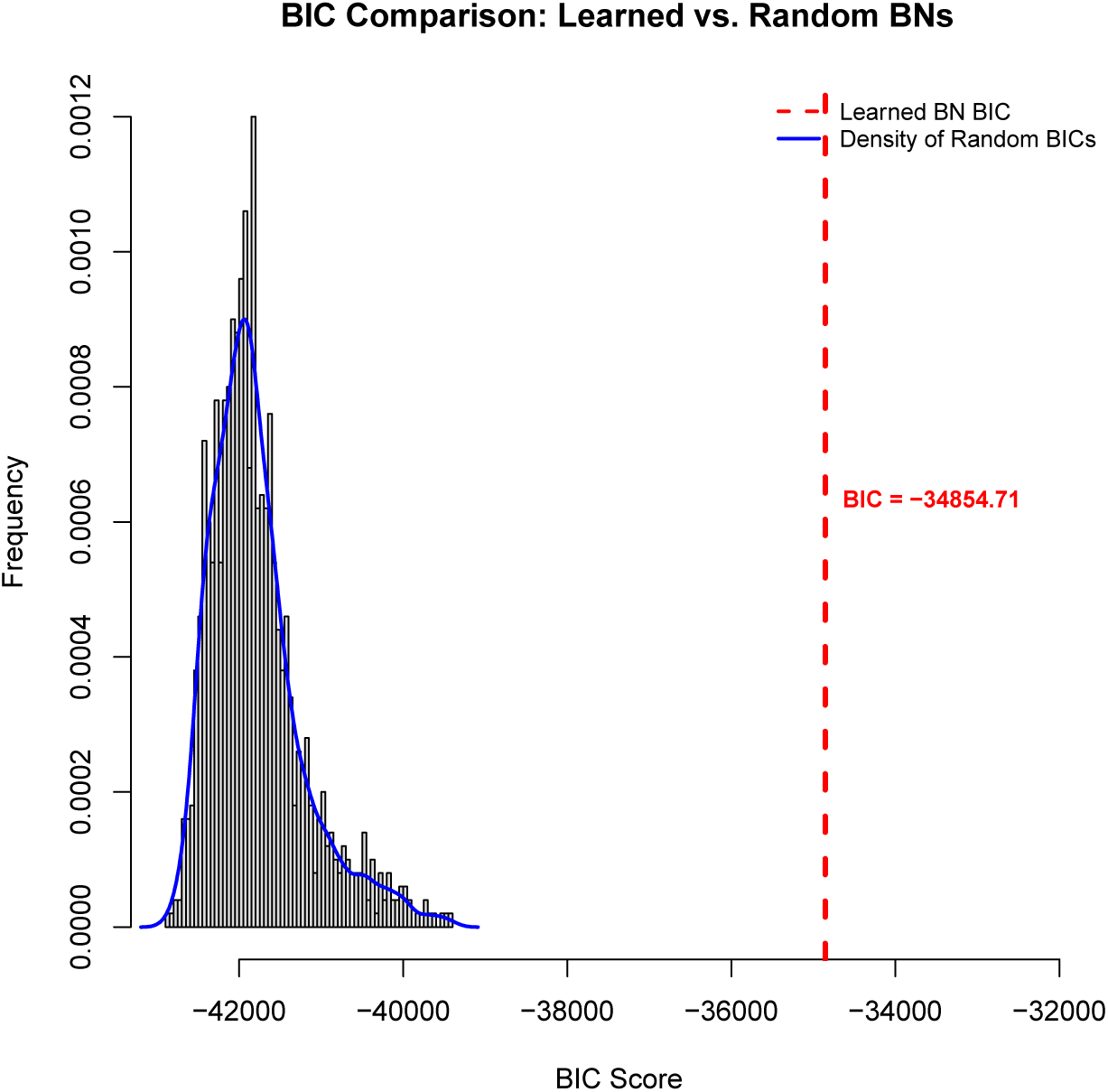
Bayesian Information Criterion (BIC) comparison for non-diabetes network model validation (n=964) Histogram of BIC scores from 1,000 randomly generated null networks (gray bars) with a smoothed density overlay (blue line). The red dashed line shows the BIC of the learned network (BIC =-34854.71), indicating significantly better fit than expected by chance.

**Figure S2.**
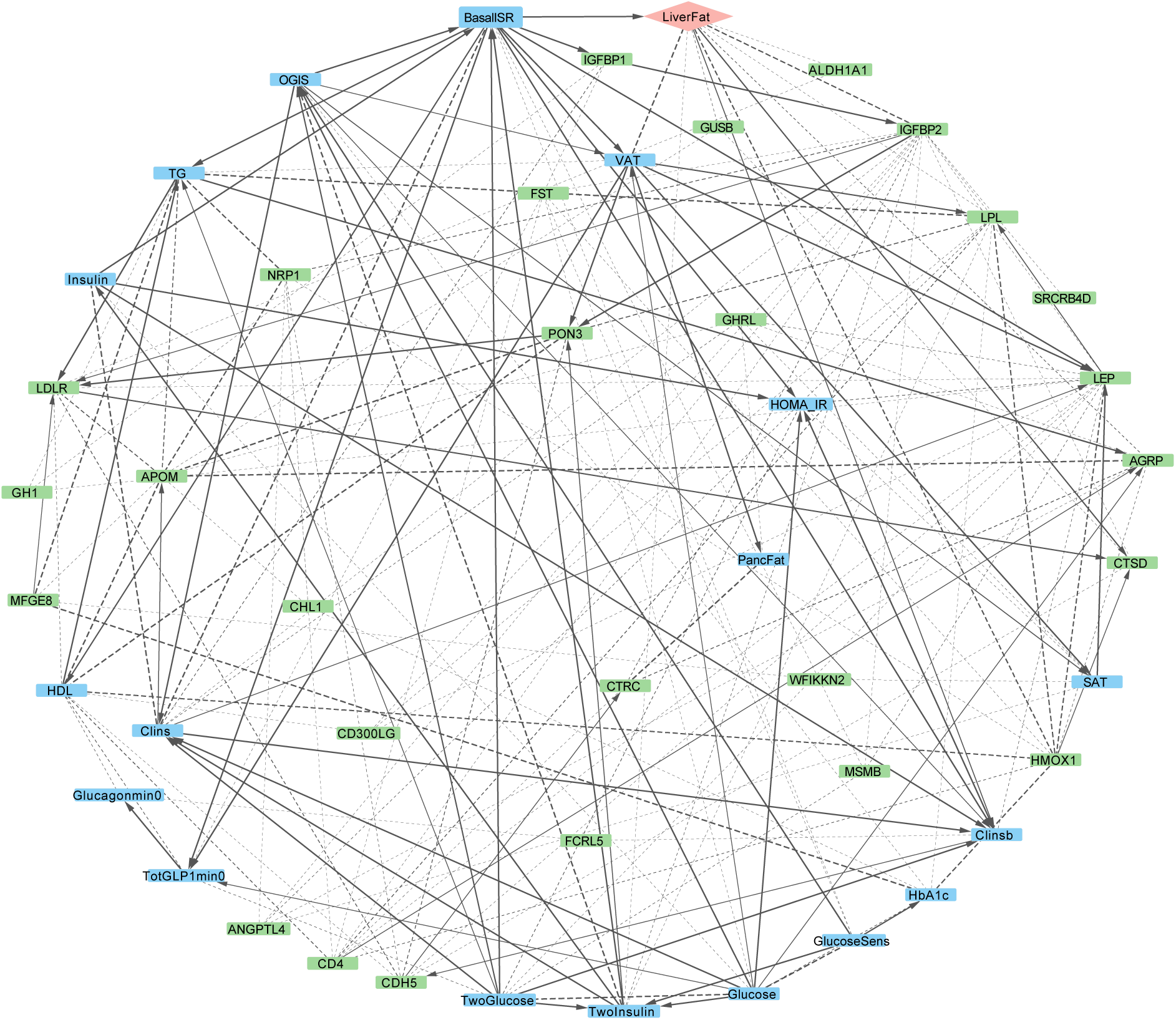
Expanded Bayesian network of metabolic and proteomic interactions in the IMI-DIRECT non-diabetes cohort (n = 964). This detailed graph shows all directed relationships (arcs with strength ≥ 0.5) among clinical and proteomic variables. Line thickness reflects the strength of association. Nodes are color-coded as follows: blue for clinical/metabolic variables, green for proteins, and peach for liver fat (outcome). Solid arrows represent directed associations with high confidence (strength and directional probability ≥ 0.8), while dashed arrows indicate less confident directionality. AGRP: agouti-related peptide; ALDH1A1: aldehyde dehydrogenase 1 family member A1; APOM: apolipoprotein M; BasalISR: basal insulin secretion rate at the beginning of the OGTT; CD4: cluster of differentiation 4; CDH5: cadherin-5; Clins: mean insulin clearance during OGTT; Clinsb: basal insulin clearance-calculated as (mean insulin secretion)/(mean insulin concentration); CTRC: chymotrypsin C; CTSD: cathepsin D; FGF21: fibroblast growth factor 21; Glucagonmin0: fasting glucagon; Glucose: fasting plasma glucose; GlucoseSens: glucose sensitivity; GUSB: β-glucuronidase; HbA1c: glycated haemoglobin A1C; HDL: high-density lipoprotein cholesterol; IGFBP1/2: insulin-like growth factor binding proteins 1 and 2; Insulin: fasting plasma insulin; KITLG: KIT ligand; LDLR: low-density lipoprotein receptor; LEP: leptin; LiverFat: hepatic fat content; LPL: lipoprotein lipase; MFGE8: milk fat globule-epidermal growth factor 8; OGIS: oral glucose insulin sensitivity index according to the method of Mari et al. [32]; PancFat: pancreas fat; PON3: paraoxonase 3; SAT: subcutaneous adipose tissue; TG: triglycerides; TotGLP1min0: fasting total GLP-1; TwoGlucose/TwoInsulin: 2-hour post-load values from OGTT; VAT: visceral adipose tissue.

**Figure S3.**
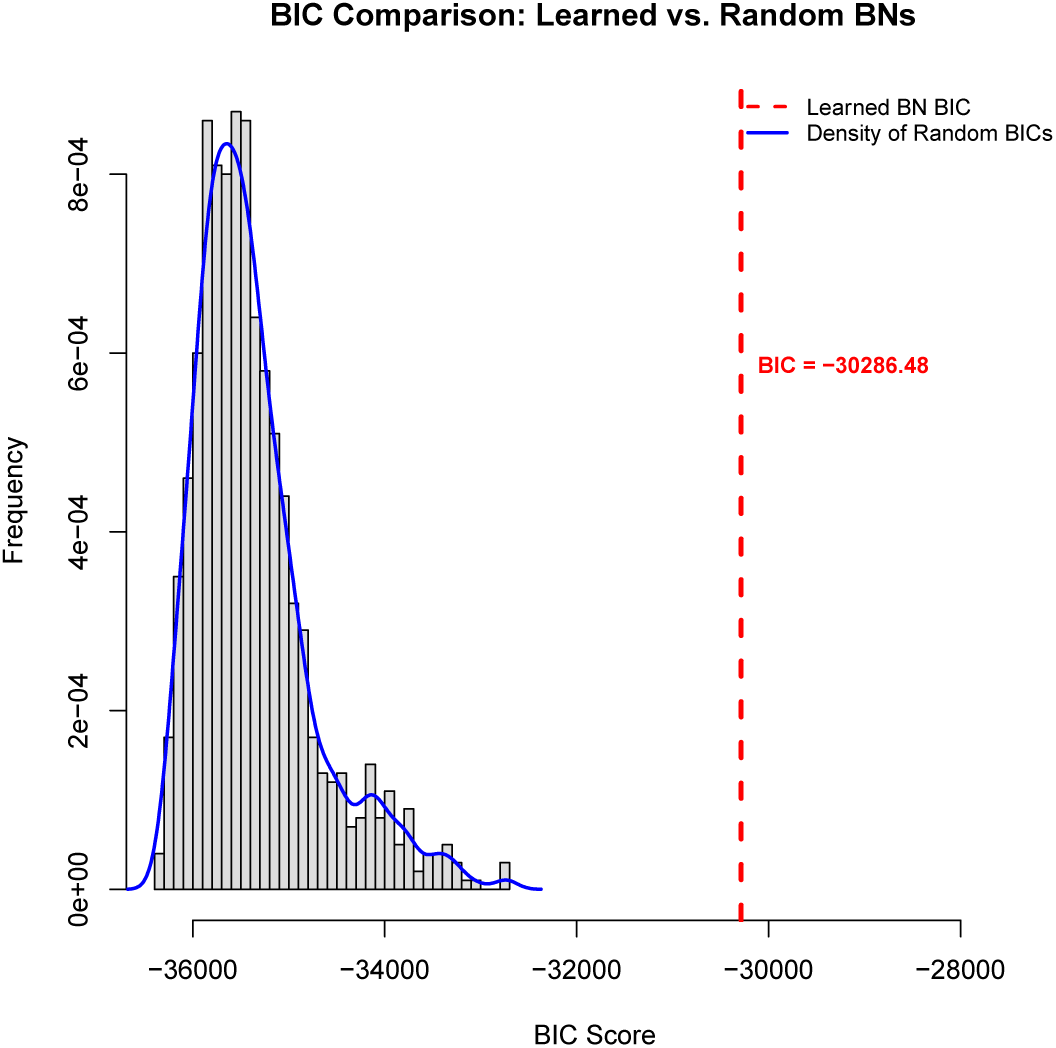
Bayesian Information Criterion (BIC) comparison for diabetes network model validation (n=331) Histogram of BIC scores from 1,000 randomly generated null networks (gray bars) with a smoothed density overlay (blue line). The red dashed line shows the BIC of the learned network (BIC =-30286.48), indicating significantly better fit than expected by chance.

**Figure S4.**
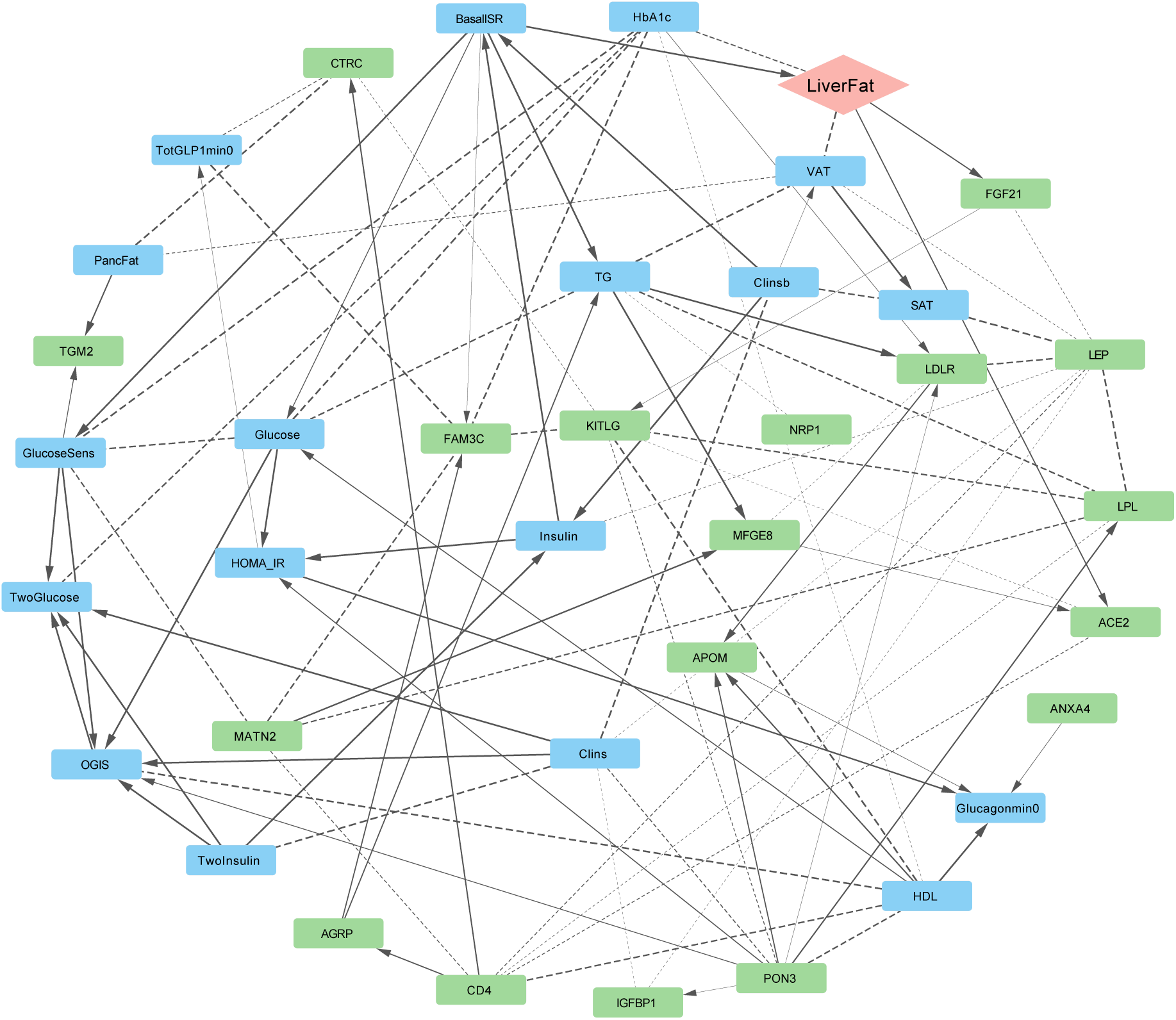
Expanded Bayesian network of metabolic and proteomic interactions in the IMI-type 2 diabetes cohort (n = 331). This detailed graph shows all directed relationships (arcs with strength ≥ 0.5) among clinical and proteomic variables. Line thickness reflects the strength of association. Nodes are color-coded as follows: blue for clinical/metabolic variables, green for proteins, and peach for liver fat (outcome). Solid arrows represent directed associations with high confidence (strength and directional probability ≥ 0.8), while dashed arrows indicate less confident directionality. ACE2: angiotensin-converting enzyme 2; AGRP: agouti-related peptide; ALDH1A1: aldehyde dehydrogenase 1 family member A1; ANXA4: annexin A4; APOM: apolipoprotein M; BasalISR: basal insulin secretion rate; CD4: cluster of differentiation 4; Clins: mean insulin clearance during OGTT; Clinsb: basal insulin clearance; CTRC: chymotrypsin C; FGF21: fibroblast growth factor 21; FAM3C: family with sequence similarity 3 member C; Glucagonmin0: fasting glucagon; Glucose: fasting plasma glucose; GlucoseSens: glucose sensitivity; HDL: high-density lipoprotein cholesterol; HOMA_IR: homeostatic model assessment of insulin resistance; IGFBP1: insulin-like growth factor binding protein 1; Insulin: fasting plasma insulin; KITLG: KIT ligand; LDLR: low-density lipoprotein receptor; LEP: leptin; LiverFat: hepatic fat content; LPL: lipoprotein lipase; MATN2: matrilin-2; MFGE8: milk fat globule-EGF factor 8; NRP1: neuropilin-1; OGIS: oral glucose insulin sensitivity index according to the method of Mari et al. [32]; PancFat: pancreas fat; PON3: paraoxonase 3; SAT: subcutaneous adipose tissue; TG: triglycerides; TGM2: transglutaminase 2; TotGLP1min0: fasting total GLP-1; TwoGlucose: 2-hour post-load glucose (OGTT); TwoInsulin: 2-hour post-load insulin (OGTT); VAT: visceral adipose tissue.

**Figure S5.**
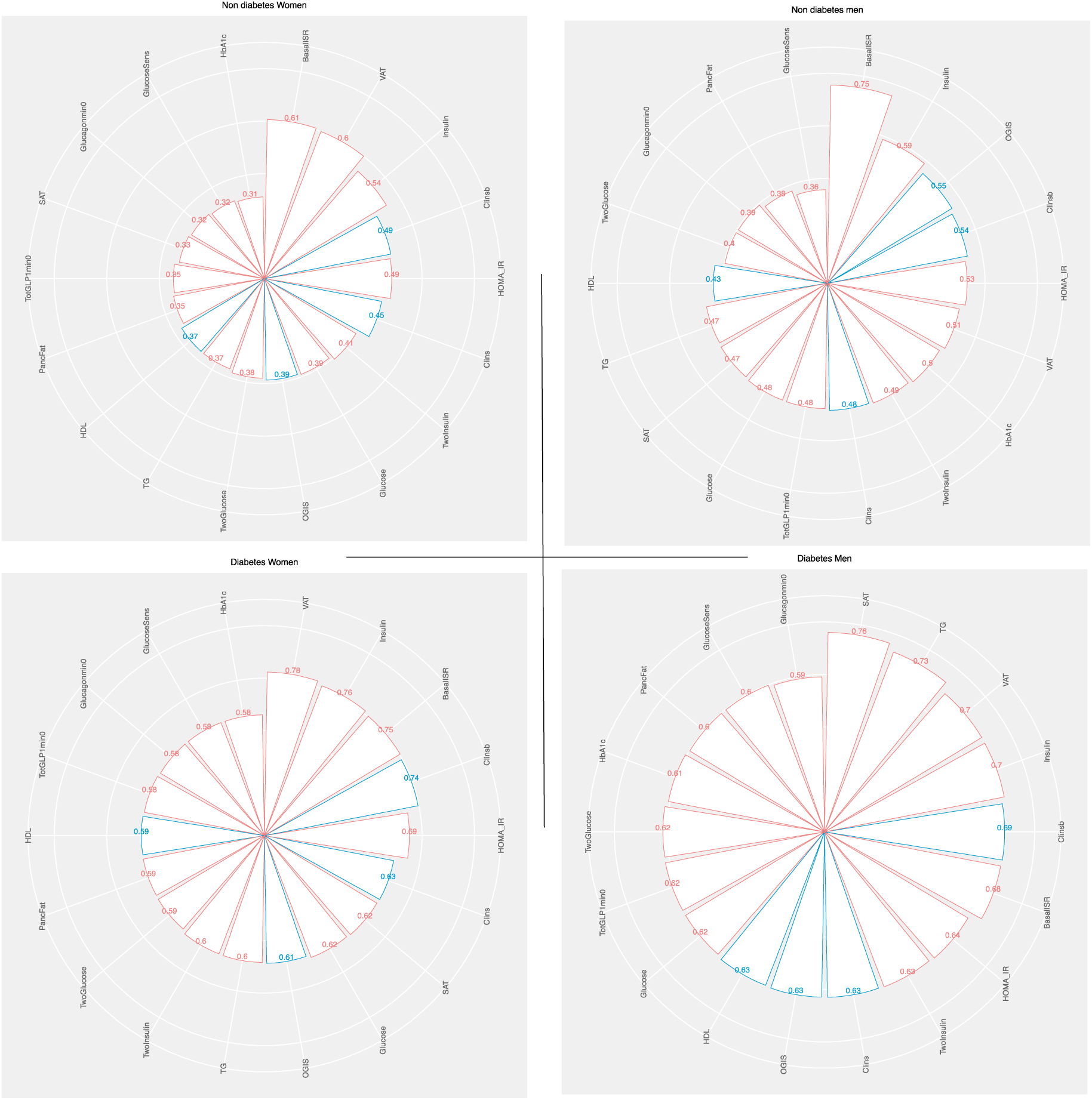
Sex-stratified Posterior probabilities of MASLD based on clinical predictors across IMI-DIRECT diabetes and non-diabetes cohorts. Radar plots display MASLD conditional probabilities following conditioning on individual clinical variables, stratified by sex and diabetes status: non-diabetic females, n=171 (top left), non-diabetic males, n=793 (top right), diabetic females, n=139 (bottom left), and diabetic males, n=193 (bottom right). Red bars indicate high levels and blue bars low levels of the respective variable. MASLD: metabolic dysfunction-associated steatotic liver disease; T2D: type 2 diabetes; BasalISR: basal insulin secretion rate; VAT: visceral adipose tissue; SAT: subcutaneous adipose tissue; OGIS: oral glucose insulin sensitivity index; HOMA_IR: homeostatic model assessment of insulin resistance; Clinsb: basal insulin clearance; Clins: dynamic insulin clearance; TG: triglycerides; HDL: high-density lipoprotein cholesterol; HbA1c: glycated haemoglobin A1c; GLP1: glucagon-like peptide 1; OGTT: oral glucose tolerance test.

**Figure S6.**
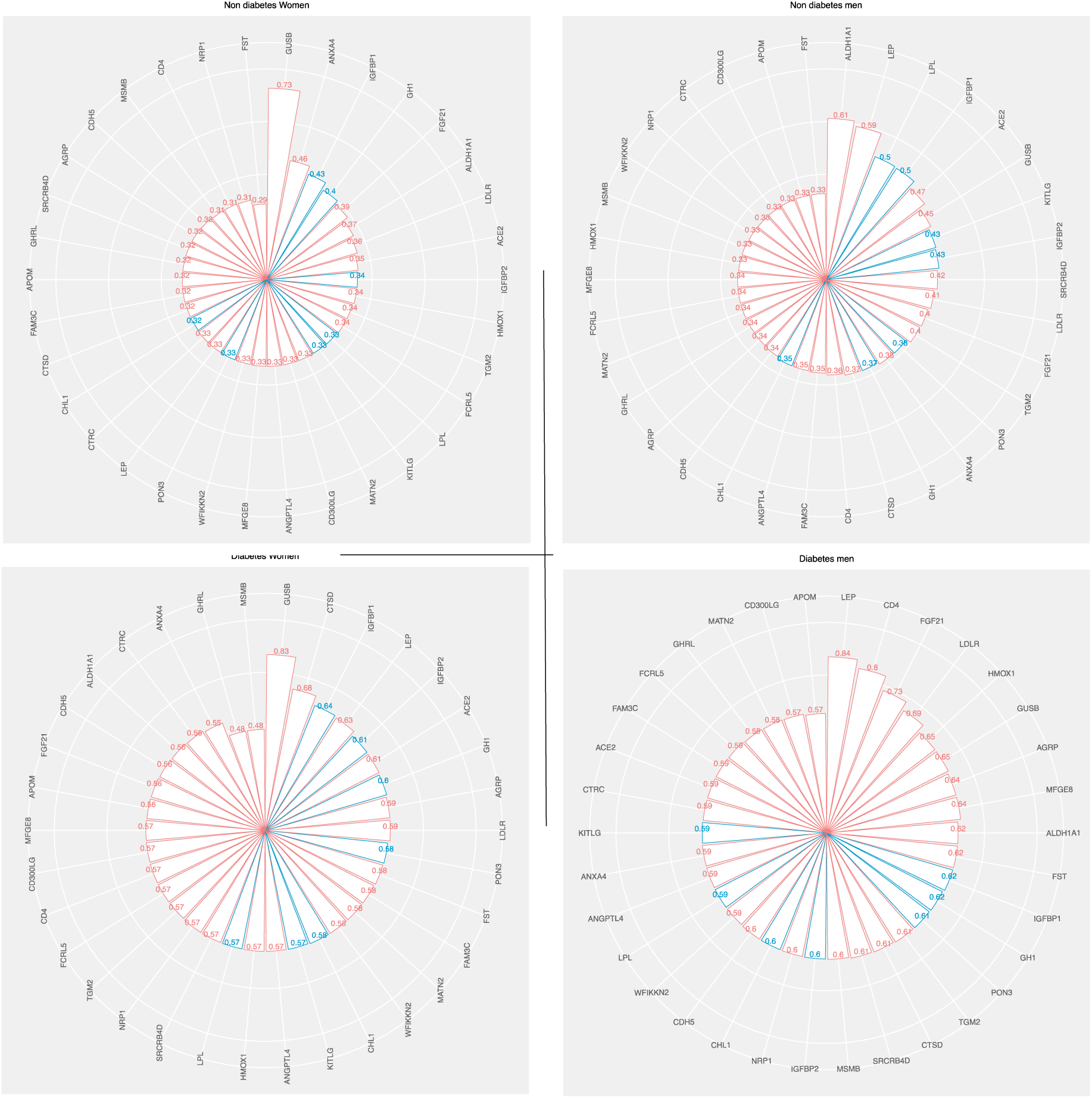
Sex-stratified Posterior probabilities of MASLD based on proteomic predictors in IMI-DIRECT diabetes and non-diabetes cohorts. Radar plots display MASLD conditional probabilities after conditioning on proteomic variables, stratified by sex and diabetes status: non-diabetic females, n=171 (top left), non-diabetic males, n=793 (top right), diabetic females, n=138 (bottom left), and diabetic males, n=193 (bottom right). Red bars show the conditional probability given high levels, and blue bars indicate conditioning on low levels of each protein. MASLD: metabolic dysfunction-associated steatotic liver disease; T2D: type 2 diabetes; GUSB: β-glucuronidase; LEP: leptin; ALDH1A1: aldehyde dehydrogenase 1 family member A1; CTSD: cathepsin D; FGF21: fibroblast growth factor 21; IGFBP1/2: insulin-like growth factor binding proteins 1 and 2; LDLR: low-density lipoprotein receptor; MFGE8: milk fat globule-EGF factor 8; AGRP: agouti-related peptide; KITLG: KIT ligand; ACE2: angiotensin-converting enzyme 2; APOM: apolipoprotein M; CD4: cluster of differentiation 4; PON3: paraoxonase 3; TGM2: transglutaminase 2; MATN2: matrilin-2.

